# Breathome discriminate Ischemic Heart Disease

**DOI:** 10.1101/2024.07.15.24310414

**Authors:** Basheer Abdullah Marzoog, Peter Chomakhidze, Daria Gognieva, Nina Vladimirovna Gagarina, Artemiy Silantyev, Alexander Suvorov, Ekaterina Fominykha, Philipp Kopylov

**Affiliations:** World-Class Research Center «Digital Biodesign and Personalized Healthcare», I.M. Sechenov First Moscow State Medical University (Sechenov University), 119991 Moscow, Russia; postal address: Russia, Moscow, 8-2 Trubetskaya street, 119991; University clinical Hospital number 1, Radiology department, I.M. Sechenov First Moscow State Medical University (Sechenov University), 119991 Moscow, Russia; postal address: Russia, Moscow, 8-2 Trubetskaya street, 119991

**Keywords:** Breathome, Metabolome, PTR-TOF-MS, VOCs, IHD, Optimizing, Bicycle ergometry

## Abstract

**Background:** Ischemic heart disease (IHD) impacts the quality of life and has the highest mortality rate in between other cardiovascular disease in the globe.

**Objectives:** IHD early diagnosis, management, and prevention remain underestimated due to the poor diagnostic and therapeutic strategies including the early prevention methods.

**Aims:** To assess the changes in the exhaled breath analysis, volatile organic compounds (VOCs), in patients with ischemic heart disease confirmed by stress computed tomography myocardial perfusion (CTP) imaging.

**Materials and methods:** A single center observational study included 80 participants from Moscow. The participants aged ≥ 40 years and given a written consent to participate in the study. Both groups, G1=31 with vs G2=49 without post stress induced myocardial perfusion defect, passed cardiologist consultation, anthropometric measurements, blood pressure and pulse rate, echocardiography, real time breathing at rest into PTR-TOF-MS-1000, cardio-ankle vascular index, performing bicycle ergometry, and immediately after performing bicycle ergometry repeating the breathing analysis into the PTR-TOF-MS-1000, and after three minutes from the second breath, repeat the breath into the PTR-TOF-MS-1000, then performing CTP. LASSO regression with nested cross-validation was used to find association between VOCs and existence of perfusion defect. Statistical processing was carried out using the R programming language v4.2 and Python v.3.10 [^R], STATISTICA, and IBM SPSS.

**Results:** The specificity 77.6 % [95 % confidence interval (CI); 0.666; 0.889], sensitivity 83.9 % [95 % CI; 0.692; 0.964], and accuracy of the diagnostic method using exhaled breath analysis, area under the curve (AUC) 83.8 % [95 % CI; 0.73655857; 0.91493173]. Whereas, the AUC of the bicycle ergometry 50.7 % [95 % CI; 0.388; 0.625], specificity 53.1 % [95 % CI; 0.392; 0.673], and sensitivity 48.4 % [95 % CI; 0.306; 0.657].

**Conclusion:** VOCs analysis appear to discriminate individuals with and without IHD with clinically acceptable diagnostic accuracy.

**Other:** The exhaled breath analysis reflects the myocardiocytes metabolomic signature and related intercellular homeostasis changes and regulation perturbances. Exhaled breath analysis poses a promise result to improve the diagnostic accuracy of the physical stress tests.

## Introduction

Incredibly, considering exhaled air as the mirror of the health of the organism is a promising approach for the future multifunctional strategy in terms of diagnosis, treatment, prevention, and evaluation of the patients prognosis [1]. Exhaled air contains a plethora of volatile organic compounds (VOCs) that demonstrate the health of the organism, including the health of the cardiovascular system [1,2]. The components of the exhaled air are variable according to the changes in the systems and organs of the body. For instance, changes in the components of exhaled air in a patient with a gastrointestinal tract pathology are different from a patient with a lung pathology. Therefore, we hypothesis that patients with cardiovascular disease have a different chemical biomarkers components in their exhaled air. At the same time, we suggest that patients with ischemic heart disease have different levels of the VOCs in their exhaled air according to the risk of death in the next 10 years using the formula of the European Society of Cardiology (SCORE2, SCORE2-OP, and SMART Risk Score) [3–6].

Various types of mass spectrometry developed in the recent years in hope of accurately analyzing exhaled breath volatile compounds. Mass spectrometry is a unique technique in which atoms and molecules of a sample are ionized, accelerated to MeV energies, and separated according to their momentum, charge, and energy, allowing high discrimination for measurement of isotope abundances [7].

Exhaled air analysis has been performed in patients with different pathologies including chronic obstructive lung disease, cancer, asthma, lung cancer, diabetes, arthritis, heart failure, gastric cancer, chronic kidney disease, colorectal cancer, hepatocellular carcinoma, malignant pleural mesothelioma, bladder cancer, pancreatic ductal adenocarcinoma, gastro-oesophageal cancer, peritonitis-shock, head and neck squamous cell carcinoma, multiple sclerosis, and Parkinson’s disease [8,9,18–27,10,28–37,11,38–47,12–17].

Despite the current advances in the technologies and therapeutic strategies, identification the origin of the molecules in the exhaled air analysis remains a challenge for scientists. The compounds of the exhaled air depend on several factors of which exogenous and endogenous. Part of the endogenous factors is the presence of pathologies in the organism, including ischemic heart disease. However, exogenous factors play critical role in the components of the exhaled air such smoking. Where, smoking is associated with 80 molecules (unsaturated hydrocarbons; 29 dienes, 27 alkenes, and 3 alkynes) in the analysis of exhaled breath compared to non-smokers [48].

Additionally, the precision and greater chance of detection of some VOCs require special preconditions including the selection of the relevant breath fraction, the type of breath collection container (if used), and the preconcentration technique [49]. Sampling of the late expiratory breath is preferred to get a greater endogenous contribution [49]. Additionally, the breath collection containers must not have a condensation effect on the collected sample. For these reasons, the scientific community requires further development a protocol in the preferred methods for collecting, processing, evaluating the results of the exhaled breath air analysis [49].

Cardiovascular disease (CVD) are the leading cause of mortality and morbidity in our era despite the current advance in therapeutic strategies and technologies [50]. Unfortunately, each second dies a person due to CVD globally [51]. Moreover, ischemic heart disease stands in the first place on the list of most frequently cause of mortality and morbidity between the cardiovascular disease. In patients with ischemic heart disease, including energy deprivation, the changes in the exhaled breath analysis are reflected in various ways, particularly through the detection of volatile organic compounds (VOCs) and other biomarkers. However, myocardiocytes activate specific signaling pathways to survive and prolong the resistant period through elevation the necrosis threshold and transforming the myocardiocyte into dormant status. Furthermore, myocardiocytes upregulate functionality of autophagy function to improve the cellular antioxidant defense system and reduce energy expenditure [52–55].

Current research on using exhaled breath analysis for diagnosis, follow-up of treatment regime, early prevention, and prognosis determination of prognosis in ischemic heart disease patients remains in the womb of development. The study sought to improve the diagnosis of ischemic heart disease during physical exertion test using PTR-TOF-1000 real-time mass spectrometry (MS).

## Materials and methods

A non-randomized, single center, mini-invasive, parallel, cross-sectional, diagnostic, observational, case-control prospective cohort study included two groups of participants at the University Clinical Hospital-1.

### Data collection

The first group consisted of 31 people with stress induced myocardial perfusion defect on the stressed computer tomography myocardial perfusion Imaging (by using contrast enhanced multi-slice spiral computed tomography (CE-MSCT) using adenosine triphosphate (ATP) as a stress test).

The second group included 49 people without stress induced myocardial perfusion defect on the stress computed tomography myocardial perfusion imaging (by using contrast enhanced multi-slice spiral computed tomography (CE-MSCT) using adenosine triphosphate (ATP) as a stress test). Additionally, the health of the participants with be confirmed by the medical history, previous medical analyses, and retrospective interviews. The study included both males and females, and the age of the participants ≥ 40 years. All the participants assessed their anthropometric measurements, blood pressure and pulse rate before starting the study, and at rest.

The study evaluated continuous and categorical variables. The continuous variables included; age, pulse at rest, systolic blood pressure (SBP) at rest, diastolic blood pressure (DBP) at rest, body weight, height, maximum heart rate (HR) on physical stress test, watt (WT) on physical stress test, metabolic equivalent (METs) on physical stress test, reached percent on physical stress test, ejection fraction (EF %) on echocardiography, estimated vessel age, right cardio-ankle vascular index (R-CAVI), left Cardio-ankle vascular index (L-CAVI), mean CAVI (=(right-CAVI + left-CAVI)/2), right ankle-brachial index (RABI), left ankle-brachial index (LABI), mean ankle-brachial index (ABI), mean SBP brachial (SBPB) (=(right SBPB+ left SBPB)/2), mean DBPB (=(right DBPB + left DBPB)/2), BP right brachial (BPRB) (=(SBP+DBP)/2), BP left brachial (BPLB) (=(SBP+DBP)/2), mean BPB (=(BPRB+BPLB)/2), BP right ankle (BPRA) (=(SBP+DBP)/2), BP left ankle (BPLA) (=(SBP+DBP)/2), mean BPA (=(BPRA+ BPLA) /2), right brachial pulse (RTb), left brachial pulse (LTb), mean Tb (=(LTb+ RTb)/2), right brachial-ankle pulse (Tba), left brachial-ankle pulse (Tba), mean Tba (= (left Tba+right Tba)/2), length heart-ankle (Lha in cm), heart-ankle pulse wave velocity (haPWV = Lha/(mean left Tba+ mean right Tba); m/s), β-stiffness index from PWV (=2*1050*( haPWV)^2*LN((mean SBPB *133,32/ mean DBPB *133,32))/((mean SBPB*133,32)-( mean DBPB *133,32)), creatinine (µmol/L), and eGFR (2021 CKD-EPI Creatinine). Categorical variables included; gender, obesity stage, smoking, concomitant disease, coronary artery, hemodynamically significant (>60%), myocardial perfusion defect after stress ATP, myocardial perfusion defect before stress ATP, atherosclerosis in other arteries (Yes/No), carotid atherosclerosis, brachiocephalic atherosclerosis, arterial hypertension (AH), stage of the AH, degree of the AH, risk of cardiovascular disease (CVD), stable coronary artery disease (SCAD), functional class (FC) by Watt and by METs, reaction type to stress test(positive/negative), reason of discontinuation of the stress test, CAVI degree, and ABI degree.

### The selection criteria involved

The inclusion criteria;

1. Participants age ≥ 40 years;
2. Participants with intact mental and physical activity;
3. Written consent to participate in the study, take blood samples, and anonymously publish the results of the study;
4. Participants in the experimental group are individuals with coronary artery disease, confirmed by stress-induced myocardial perfusion defect on the adenosine triphosphate stress myocardial perfusion computed tomography.

Exclusion criteria:

1. Failure of the stress test for reasons unrelated to heart disease;
2. Reluctance to continue participating in the study.

Non-inclusion criteria

1. Pregnancy and breast feeding.
2. Diabetes mellitus.
3. Presence of signs of acute coronary syndrome (myocardial infarction in the last two days), history of myocardial infarction;
4. Active infectious and non-infectious inflammatory diseases in the exacerbation phase;
5. Respiratory diseases (bronchial asthma, chronic bronchitis, cystic fibrosis);
6. Acute thromboembolism of pulmonary artery branches;
7. Aortic dissection;
8. Critical anatomical heart defects;
9. Active oncopathology;
10. Decompensation phase of acute heart failure;
11. Neurological pathology (Parkinson’s disease, multiple sclerosis, acute psychosis, Guillain-Barré syndrome);
12. Cardiac arrhythmias that do not allow exercise ECG testing (Wolff-Parkinson-White syndrome, Sick sinus syndrome, AV block of II-III-degree, persistent ventricular tachycardia);
13. Diseases of the musculoskeletal system that prevent passing a stress test (bicycle ergometry);
14. Allergic reaction to iodine and/or adenosine triphosphate.

The current paper is a PhD work by MD. Basheer A. Marzoog. The study registered at the clinicaltrails.gov (NCT06181799), and the study approved by the Sechenov University, Russia, from “Ethics Committee Requirement № 19-23 from 26.10.2023”. A written consent is taken from the study participants for publication of any obtained results including figures.

### Vessel stiffness measurement

Both groups passed a vessel stiffness test and pulse wave recording as well as vascular age by using Fukuda Denshi device (VaSera VS-1500; Japan). Cuffs placed to assess the vascular stiffness and the vascular age as well as the ancle-brachial index.

Cuffs fit to the size of the arms and ankles of the patients. Electrodes attach to the two arms, and a microphone for cardio-phonogram measurements fix with double-sided tape over the sternum in the second intercostal space. Cardio-ankle vascular index (CAVI parameter) reflects the overall stiffness of the aorta, femoral artery and tibial artery, and is theoretically not affected by blood pressure [56]. CAVI measurements considered valid only when obtained during at least three consecutive heartbeats [56]. These CAVI measurements to exclude vascular pathology and determine the biological age of the blood vessels. The measurement of the vascular stiffness and estimated vascular ae is to determine the state of the non-coronary arteries.

### Physical exertion test

Subsequently, participants passed exercise bicycle ergometry (on SCHILLER CS200 device; Bruce protocol or modified Bruce protocol) test to evaluate the response to physical activity. And just after completing the exercise test, the participants exhaled a second time into the same real time mass spectrometry, within one minute. And a third time exhaled into the same mass spectrometry after three minutes from the end of the second breathing, within one minute. According to the results metabolic equivalent; Mets-ВT (ВТ), the angina functional class (FC) in participants with positive stress test results determined, Where ВТ/Mets <50/<4 FC-III, ВТ/Mets 50-100/4-7 FC-II, ВТ/Mets >100/7 FC-I. During the bicycle ergometry test, the participants monitored with 12-lead ECG and manual blood pressure measurement, 1 time each 2 minutes, close to the end of each stage.

The ergometry procedure discontinued, if an increase in systolic blood pressure ≥ 220 mmHg or horizontal or downsloping ST segment depression on the ECG ≥ 1 mm, typical heart pain during test, ventricular tachycardia or atrial fibrillation, or other significant heart rhythm disorders were found Moreover, stop the procedure if the target heart rate (≥ 86% of the 220-age) is reached.

### Stressed computer tomography myocardial perfusion (CTP) imaging

Before performing the stressed computer tomography with myocardial perfusion imaging, all the participants present results of the venous creatinine level, eGFR (estimated glomerular filtration rate) according to the 2021 CKD-EPI creatinine > 30 ml/min/1,73 m2, according to the recommendation for using this formula by the National kidney foundation and the American Society of Nephrology [57–60].

The participants of both groups got catheterization in the basilar vein or the radial vein for injection of contrast and Natrii adenosine triphosphate (10 mg/1ml) to induce pharmacological stress test to the heart by increasing heart rate. Then using the catheter for contrast injection during the procedure of the computer tomography.

To prepare the Natrii Adenosine Triphosphate, a 3ml of Adenosine Triphosphate dilute in 17 ml of isotonic Sodium Chloride solution 0.9%, the injected volume of the diluted drug in milliliters is calculated by body weight. For 1 dose, take 3 ml of adenosine triphosphate (3 ampoules of each 1 ml (10mg)) + 17 ml of isotonic solution of sodium chloride 0.9% in one syringe, 20 ml. For one patient, manually inject intravenously (IV) through the already inserted catheter at a rate of 300 μg/kg/2 minutes, depending on weight: 60 kg = 12 ml, 70 kg = 14 ml, 80 kg = 16 ml, 100 kg = 20 ml of the full dose.

Stress computed tomography myocardial perfusion (CTP) imaging (done on Canon device with 640 slice, 0,5mm thickness) with contrast (Omnipaque, 50 ml). Firstly, make image to evaluate the calcification level in the valves and the ascending aorta. Then, inject the contrast and make a rest image for myocardial perfusion, then the patient continues lying on the apparat for 20 minutes and then inject in to the catheter the Natrii Adenosine Triphosphate (10 mg/1ml) according to body weight to cause pharmacological stress test to the heart during two minutes. Then make and image of the myocardial perfusion after stress test immediately, the image must be done in less than 30 seconds. (*Figure 1*)

**Figure 1:**
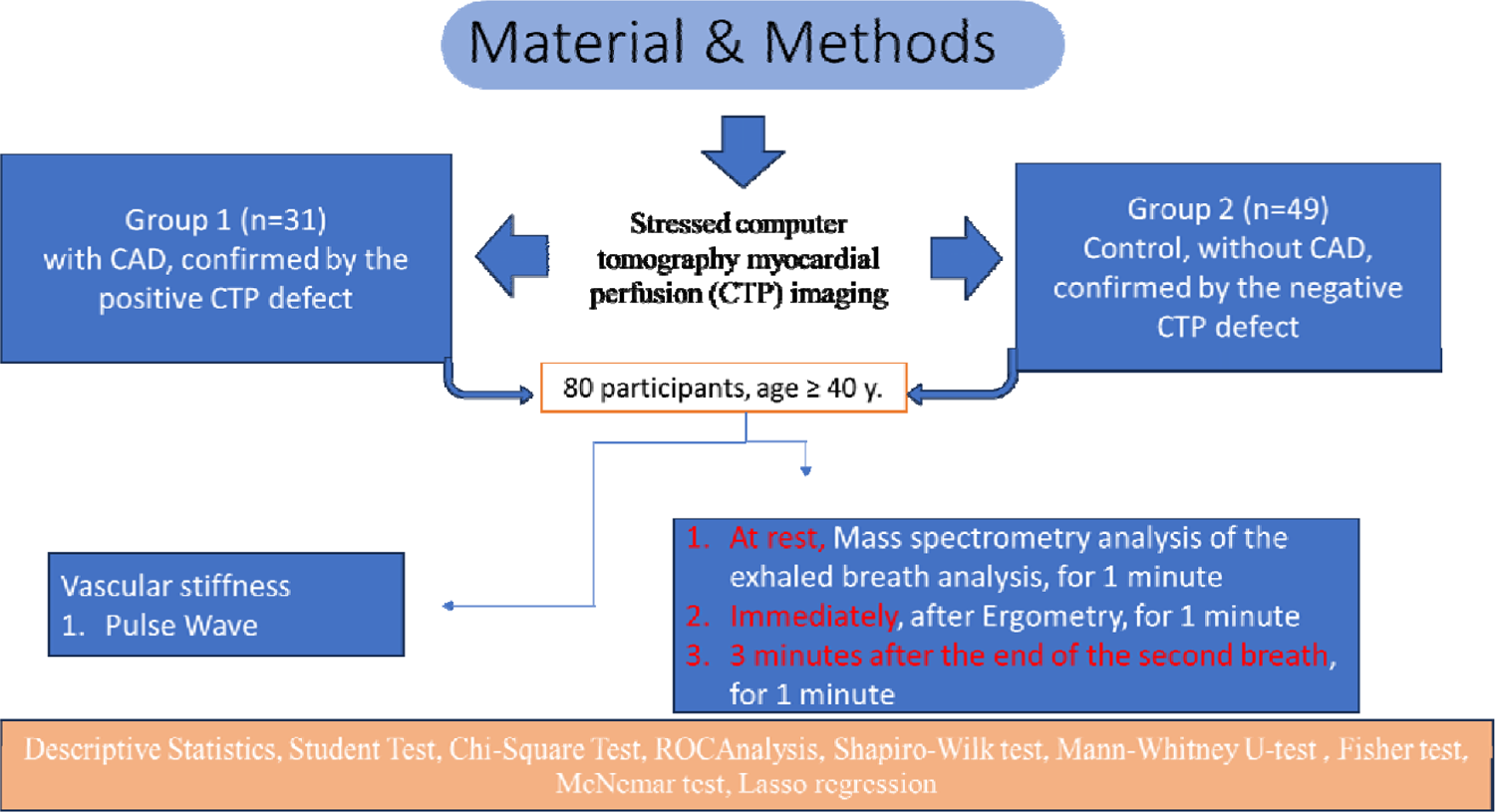
Diagrammatic presentation of the study. The patients exhale in mass spectrometer (PTR TOF-1000 (IONICON PTR-TOF-MS - Trace VOC Analyzer). Subsequently, participants pass exercise bicycle ergometry (on SCHILLER device; Bruce protocol or modified Bruce protocol). Then breath immediately after the end of the test (second breath), and after three minutes from the end of the second breath, the participants again exhale in the same mass spectrometer using single time tube.

### Mass spectrometry

All participants, at rest, passed real time mass spectrometry (MS) within one minute using a PTR TOF-1000 (IONICON PTR-TOF-MS - Trace VOC Analyzer, Eduard-Bodem-Gasse 3, 6020 Innsbruck, Austria (Europe). The analysis of exhaled air was carried out in the hospital at morning, on an empty stomach, without toothbrushing. All participants abstained from food and liquids (except water) and exercise training for 6-8 hours before breathing [61]. Participants used disposable and sterile mouthpieces, and according to the manufacturer’s instructions, additional filters were not required. All participants breathed into the sampler for 1 minute (during this time from 12 to 16 exhalation cycles are analyzed). The ionized molecules were separated by their m/z and subsequently detected. Full scan mass spectra were obtained in the 10-685 mass-to-charge ratio (m/z) with a scan time of 1000 ms and primary ion H3O+. The temperature of T-Drift and T-Inlet was 80 °

### Statistical analysis

For quantitative parameters, the nature of the distribution (using the Shapiro-Wilk test), the mean, the standard deviation, the median, the interquartile, the minimum and maximum values were determined. For categorical and qualitative features, the proportion and absolute number of values were determined.

Comparative analysis for normally distributed quantitative traits was carried out on the basis of Welch’s t-test (2 groups); for abnormally distributed quantitative traits, using the Mann-Whitney U-test (2 groups).

Comparative analysis of categorical and qualitative features was carried out using the Pearson X-square criterion, in case of its inapplicability, using the exact Fisher test.

For exhaled air values, baseline values (prefixed with “ *l0*_” were used, and deltas between immediately after exertion (l1) and after 2nd exhalation, as well as between after 2nd exhalation and immediately after exertion, were calculated:

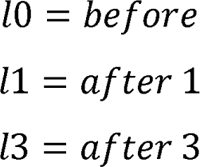

Calculation of delts:

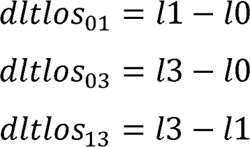

Statistical processing carried out using the R programming language v4.2, Python v.3.10 [^R], and Statistica 12 programme. (StatSoft, Inc. (2014). STATISTICA (data analysis software system), version 12. www.statsoft.com.). P considered statistically significant at <0.05.

### Outcome and feature selection with cross-validation

Due to the small number of observations (n = 80), random sampling of 2/3 of the available sample for predictor selection was performed for 1000 repetitions to evaluate the performance of the predictors. Data preprocessing at each iteration involved normalization and iterative imputation using Bayesian ridge regression for quantitative data. There were no categorical or binary features. At each iteration, a classifier was built using the gradient boosting algorithm, which made it possible to calculate feature importances for 1000 times. Then, feature importances medians were calculated for each factor, and predictors were ranked from the highest median values to the lowest.

Ten selected predictors were included in a new pipeline, the same data preprocessing was performed, then a classifier was built using the gradient boosting algorithm. Leave-one-out cross-validation was used. After that, the area under the curve, AUC, was calculated, and the optimal threshold was selected for calculating sensitivity and specificity, positive and negative prognostic values. The obtained area under the curve was compared with the result of stress test using the McNemar criterion. This procedure was performed separately exhaled breath data without the other clinical data.

## Results

### The cohort

The primary included number in the study is 101 individual, excluded 21 (either discontinued in the study or excluded due to the detection an exclusion criteria). The prospective study involved 80 participants. According to the results of the CTP, the participates divided in to two groups. The first group participants with stress induced myocardial perfusion defect (n=31) and the second group without stress induced myocardial perfusion defect (n=49) on the CTP.

### Descriptive statistics results

The descriptive characteristics of the sample were shown as both groups and then each group separately in tables for a full representation of the results. The characteristics of the continuous variables of the sample described in the below tables. (Table 1A-B)

**Table 1:** The features (Descriptive Statistics) of the continues variables of the whole sample represented in the table.

| Variable | Mean | Minimum | Maximum | Std.Dev. |
| --- | --- | --- | --- | --- |
| Age | 56.28 | 40.24 | 77.94 | 10.601 |
| Pulse rate at rest | 70.29 | 49.00 | 93.00 | 9.559 |
| SBP at rest | 123.16 | 54.00 | 159.00 | 15.437 |
| DBP at rest | 80.61 | 60.00 | 122.00 | 11.238 |
| Body weight | 77.92 | 52.50 | 140.00 | 16.236 |
| Height | 169.95 | 148.00 | 190.00 | 8.835 |
| BMI | 26.93 | 18.49 | 48.44 | 4.901 |
| Pulse rest (Cardio-Qvark®) | 69.68 | 49.00 | 99.00 | 9.260 |
| Pulse after stress (Cardio-Qvark®) | 86.89 | 63.00 | 115.00 | 10.579 |
| Goal heart rate | 163.72 | 142.06 | 179.76 | 10.601 |
| Max HR | 146.25 | 108.00 | 199.00 | 14.111 |
| Reached % | 89.53 | 64.89 | 135.25 | 9.166 |
| WT | 125.63 | 75.00 | 250.00 | 44.111 |
| METs | 6.67 | 2.90 | 11.90 | 1.977 |
| EF (%) | 64.52 | 55.00 | 73.00 | 4.388 |
| eVessel age | 56.50 | 20.00 | 80.00 | 13.520 |
| R-CAVI | 8.21 | 4.80 | 15.10 | 1.379 |
| L-CAVI | 8.18 | 4.80 | 14.90 | 1.299 |
| Mean CAVI | 8,194 | 4,800 | 15,000 | 1,331 |
| RABI | 1.15 | 0.86 | 1.40 | 0.088 |
| LABI | 1.15 | 0.89 | 1.42 | 0.084 |
| Mean SBP B | 134.38 | 105.50 | 169.00 | 13.086 |
| Mean DBP B | 85.28 | 60.50 | 104.50 | 8.358 |
| BP RB $(=(SBP+DBP)/2)$ | 103.98 | 75.00 | 137.00 | 11.547 |
| BP LB $(=(SBP+DBP)/2)$ | 104.54 | 71.00 | 136.00 | 10.534 |
| Mean BP B | 104.26 | 73.00 | 136.50 | 10.772 |
| BP RA $(=(SBP+DBP)/2)$ | 108.39 | 80.00 | 137.00 | 12.553 |
| BP LA $(=(SBP+DBP)/2)$ | 108.63 | 81.00 | 138.00 | 11.519 |
| Mean BP A | 108.51 | 81.50 | 135.00 | 11.468 |
| Mean ABI | 1.15 | 0.88 | 1.41 | 0.081 |
| RTb | 80.53 | 59.00 | 152.00 | 13.441 |
| LTb | 77.26 | 58.00 | 128.00 | 14.012 |
| Mean Tb | 78.89 | 59.00 | 132.50 | 12.547 |
| Right Tba | 86.26 | 23.00 | 117.00 | 16.232 |
| Left Tba | 86.40 | 24.00 | 114.00 | 14.685 |
| Mean Tba | 86.33 | 23.50 | 115.00 | 15.358 |
| Lha (cm) | 148.17 | 130.33 | 164.47 | 7.182 |
| haPWV (m/s) | 0.91 | 0.67 | 1.42 | 0.124 |
| β-stiffness index from PWV | 2.83 | 1.21 | 7.04 | 0.813 |
| Creatinine ($\mu\text{mol/L}$) | 82.74 | 53.90 | 138.00 | 16.014 |
| eGFR (2021 CKD-EPI Creatinine) | 85.31 | 45.40 | 113.70 | 14.684 |

**Table 1B:**
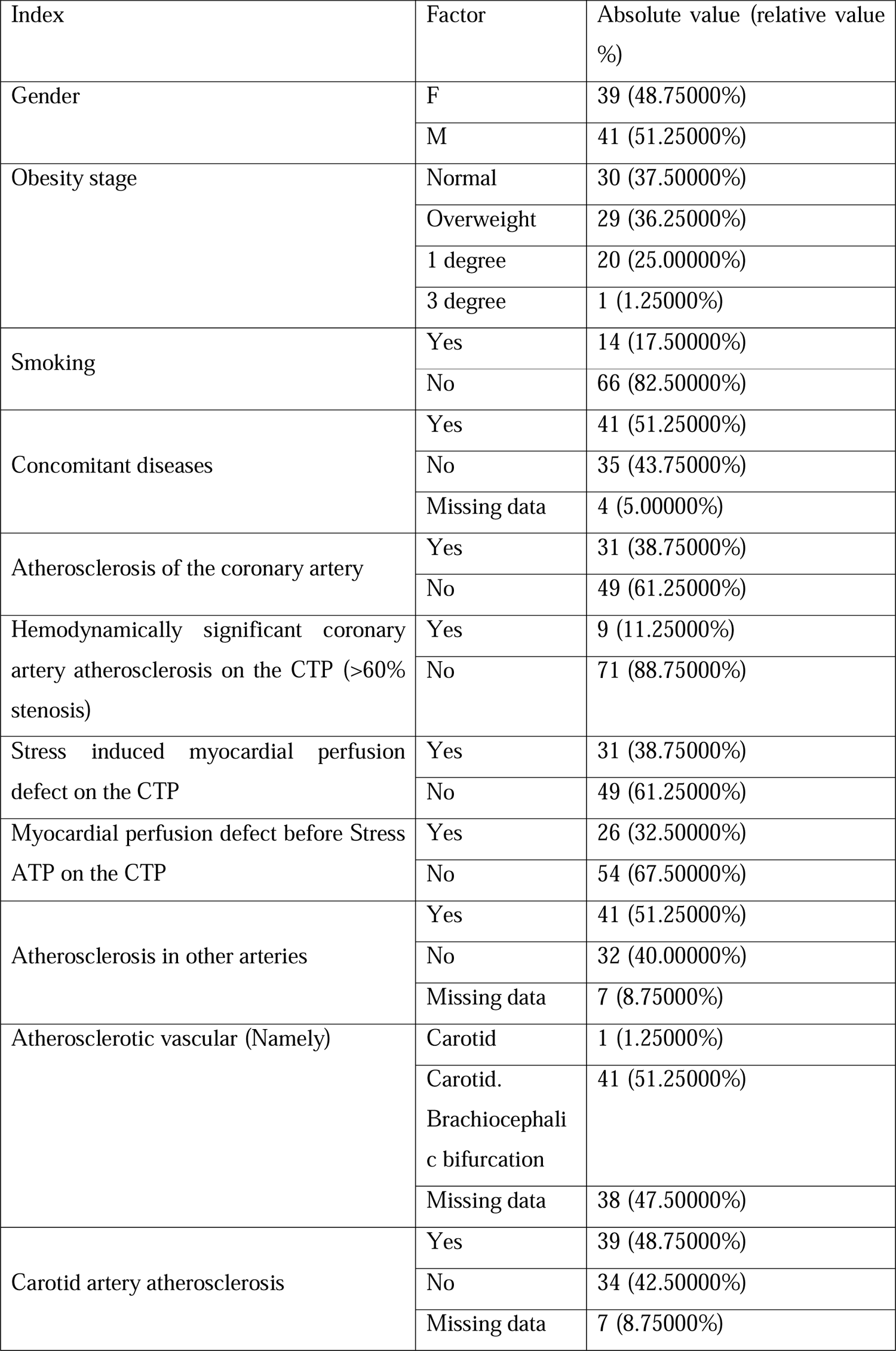
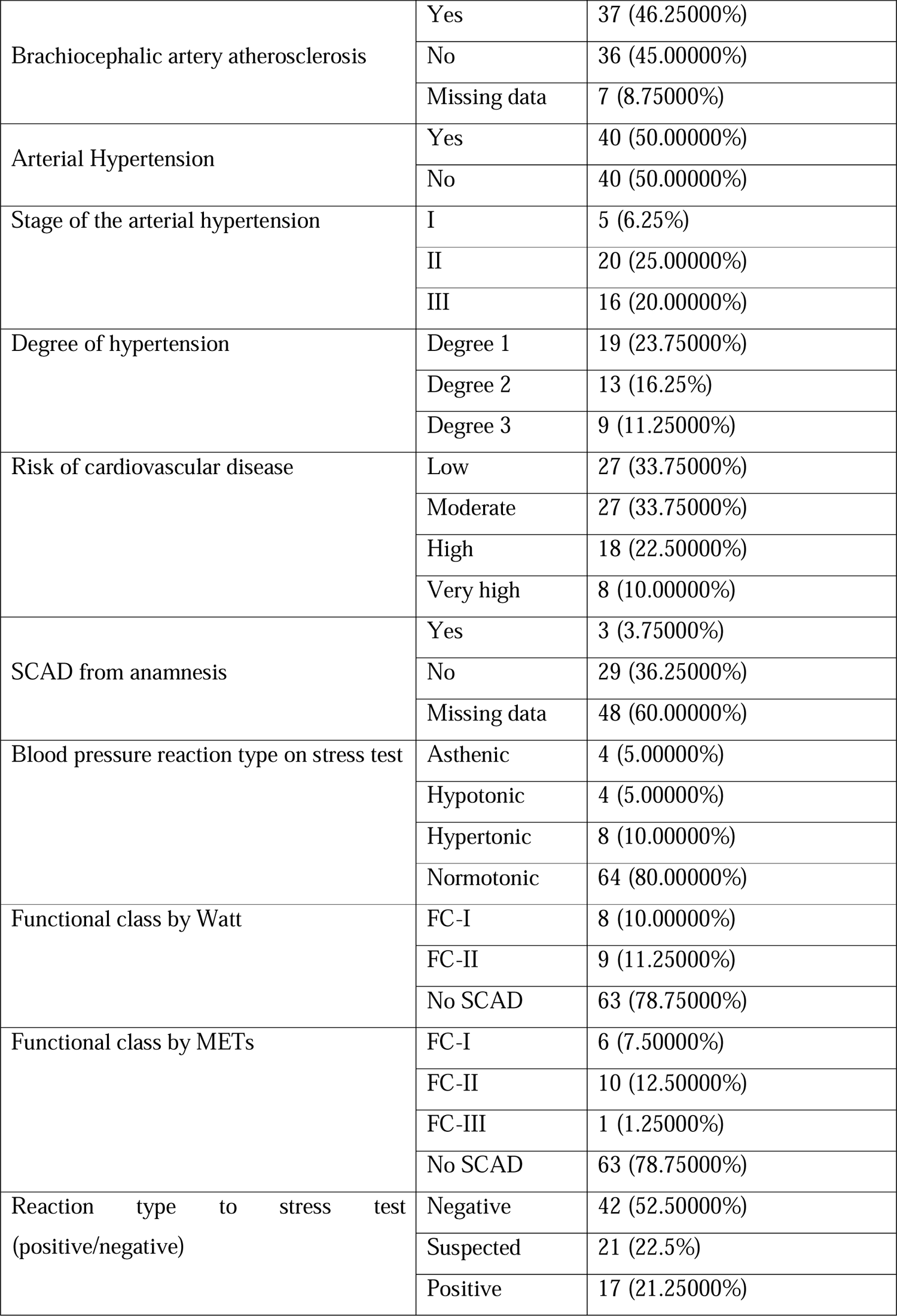
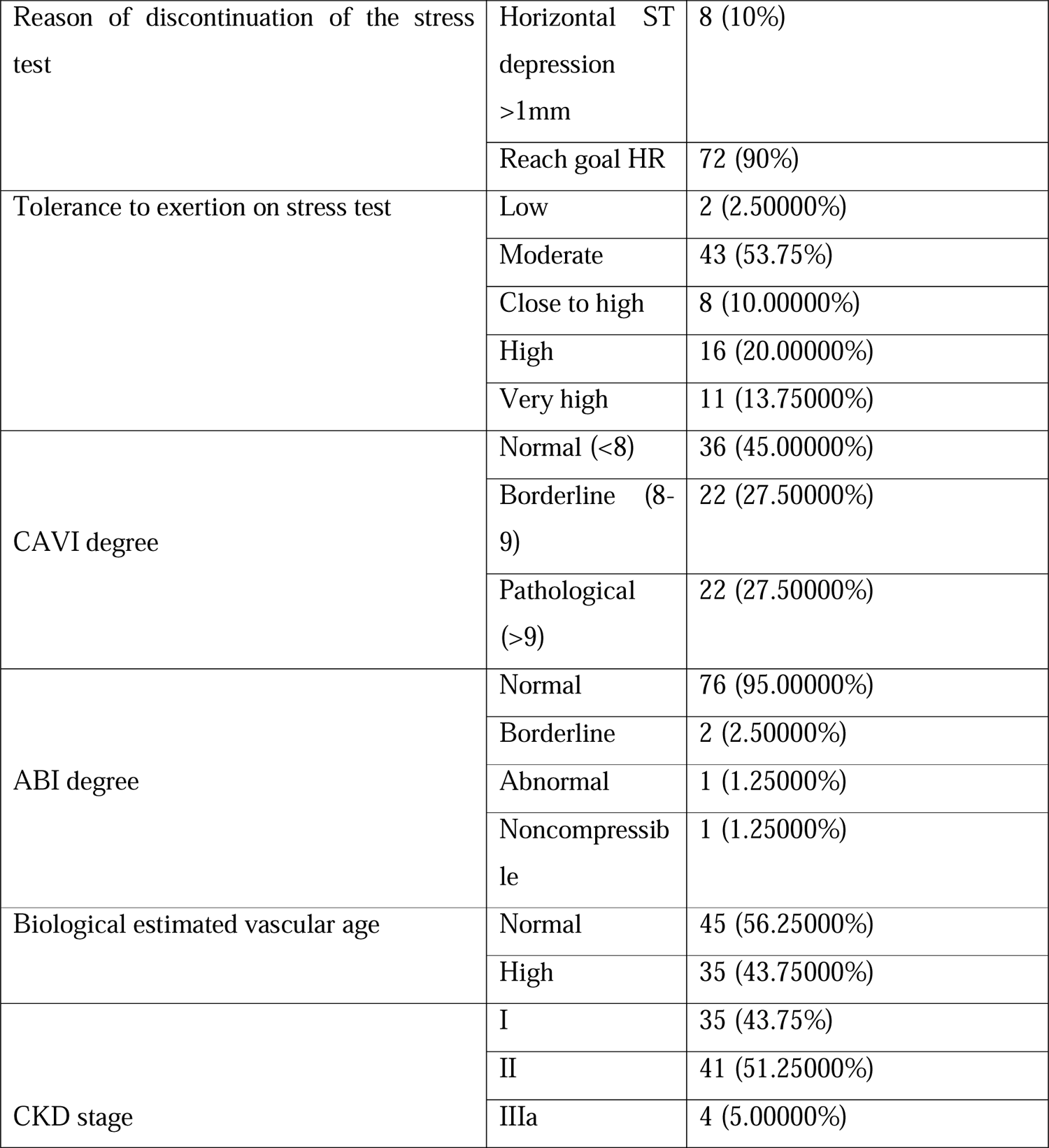
The features (Descriptive Statistics) of the categorical variables of the whole sample represented in the table. Abbreviations: METs; metabolic equivalent. CPT; stress myocardial perfusion computer tomography imaging.

The comparative characteristics of the sample represented in the below tables based on the presence or absence of the stress induced myocardial perfusion defect of the CTP imaging with the adenosine triphosphate. (*Table 2A-B*)

**Table 2A:**
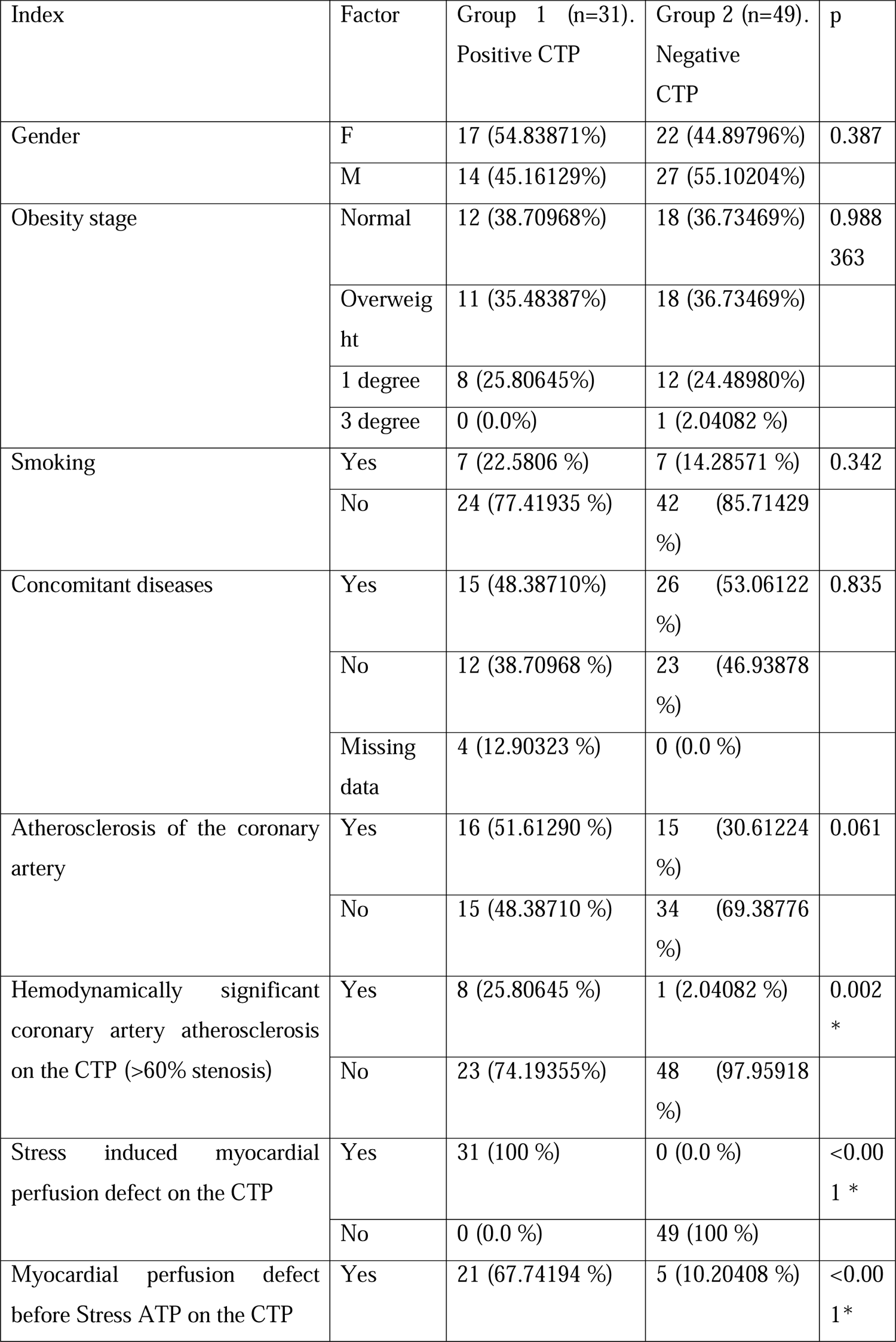
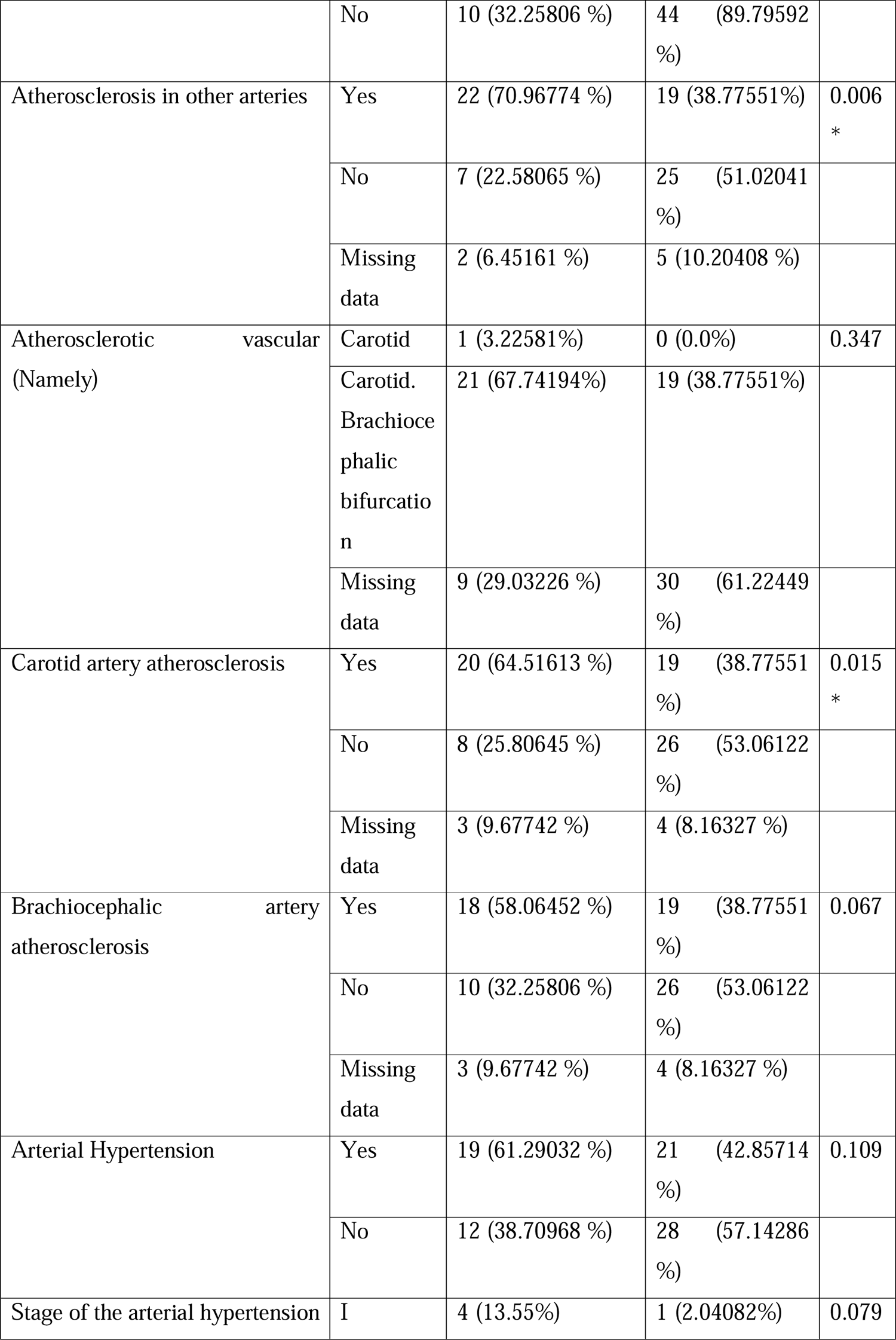
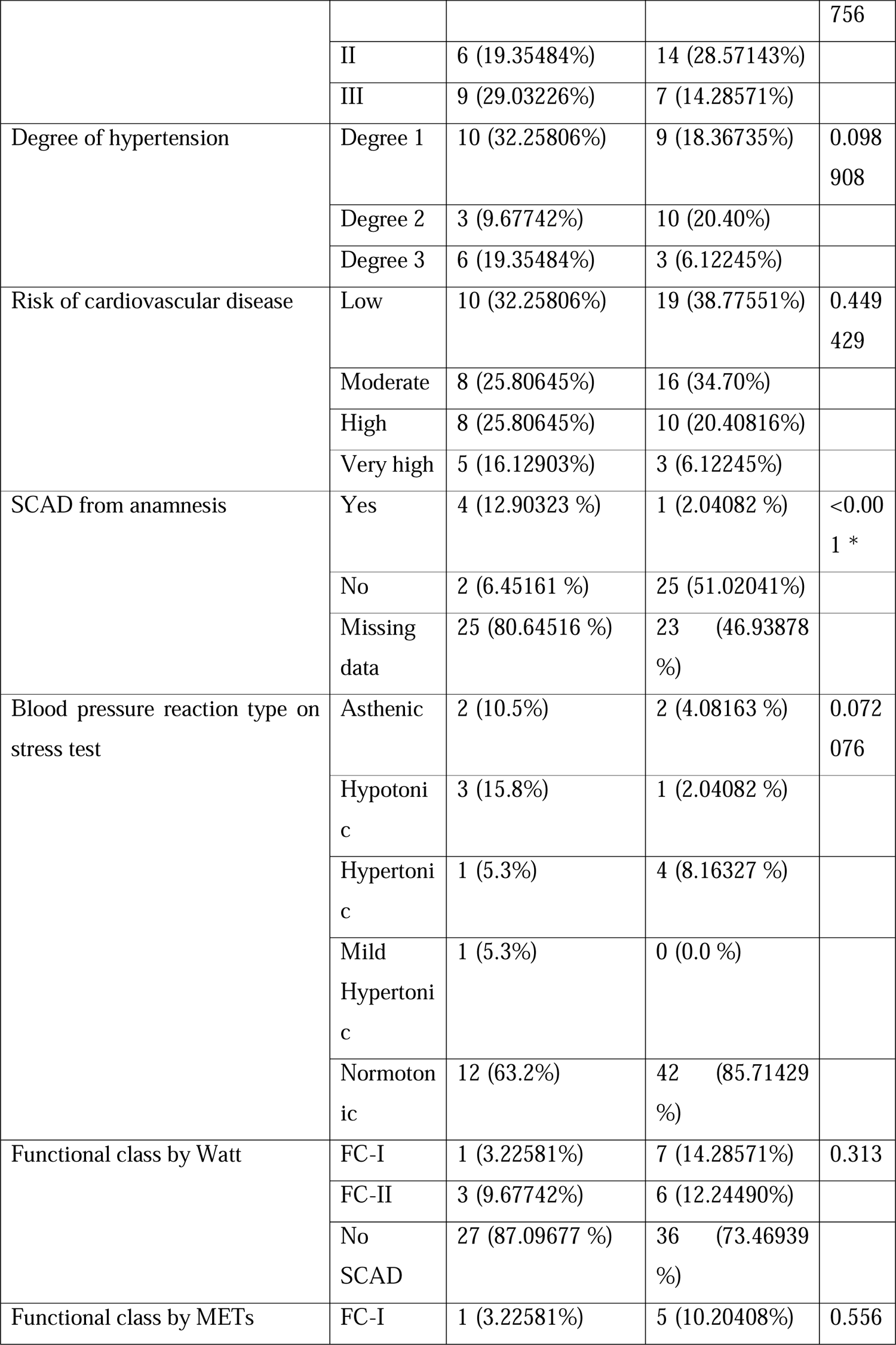
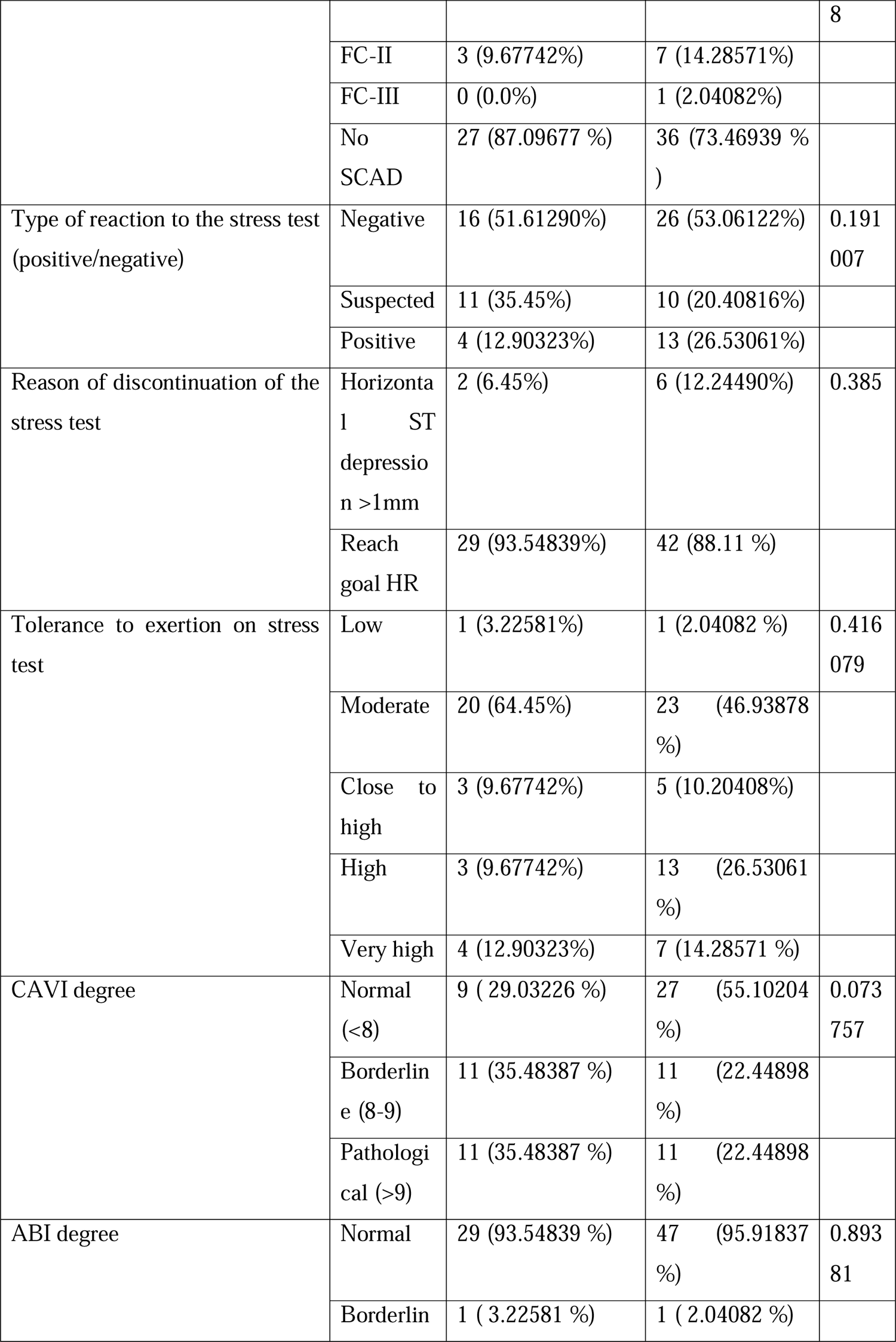
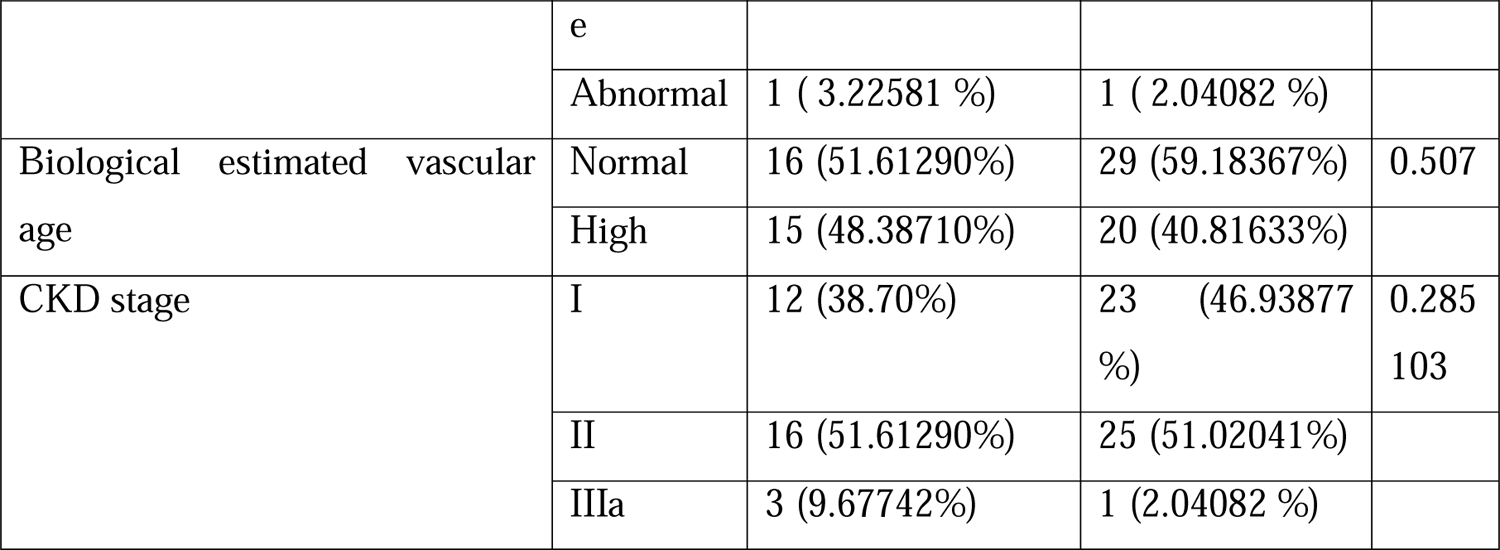
Categorical variables presented in absolute and relative values of the study for true incidence of the stated factor. X^2^ test used as a comparative test. * Values statically significant difference. Abbreviations: METs; metabolic equivalent. CPT; stress myocardial perfusion computer tomography imaging.

**Table 2B:**
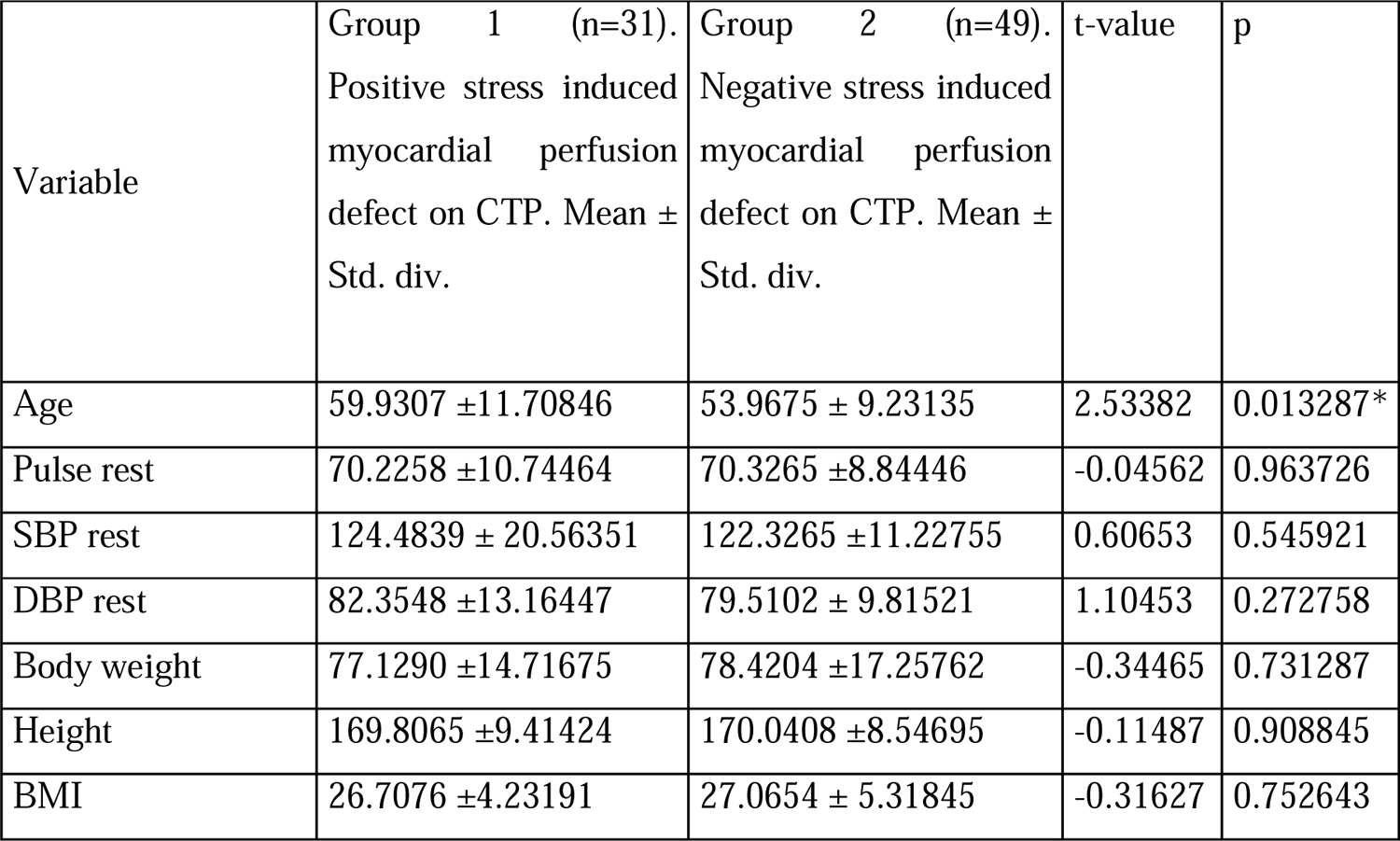
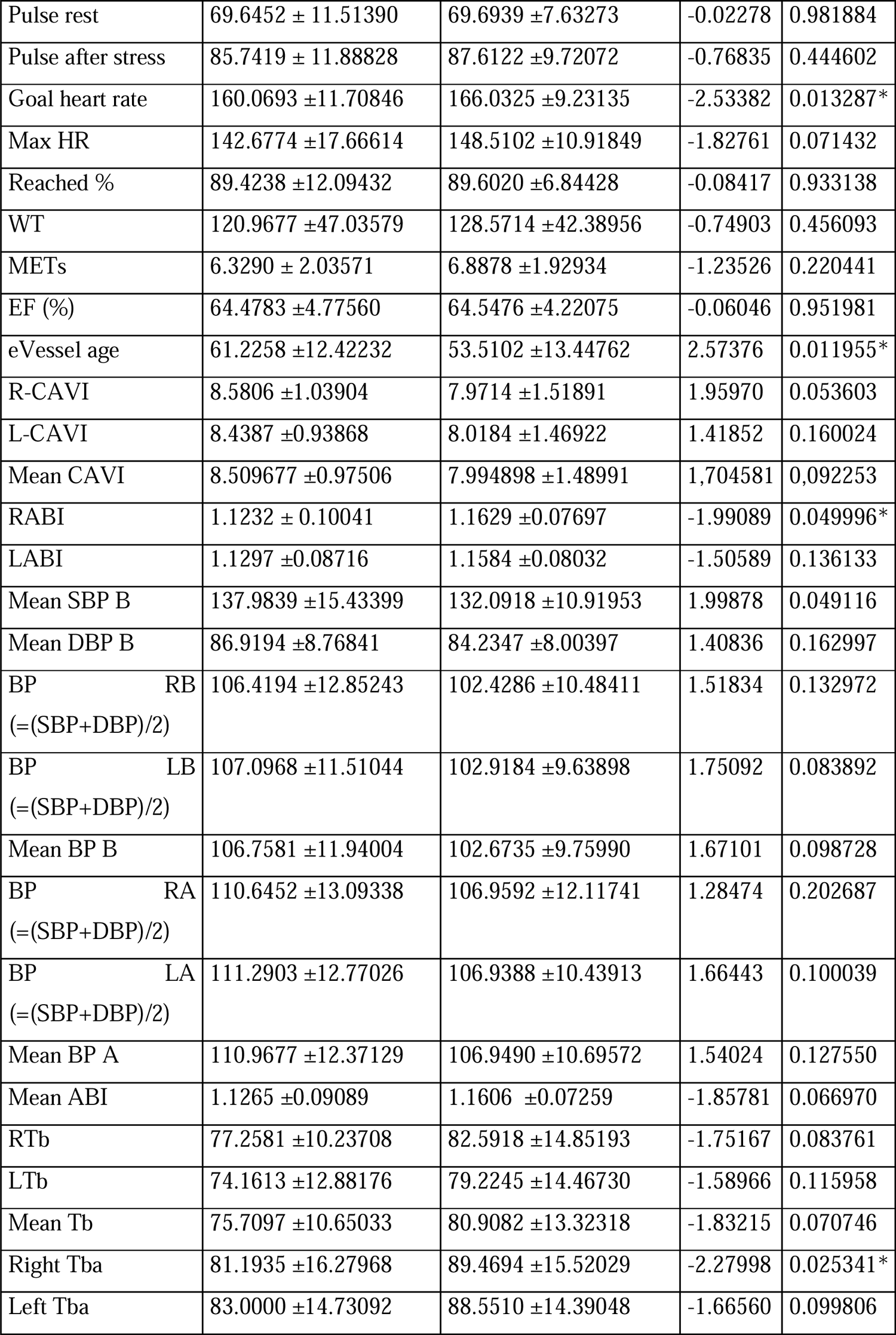
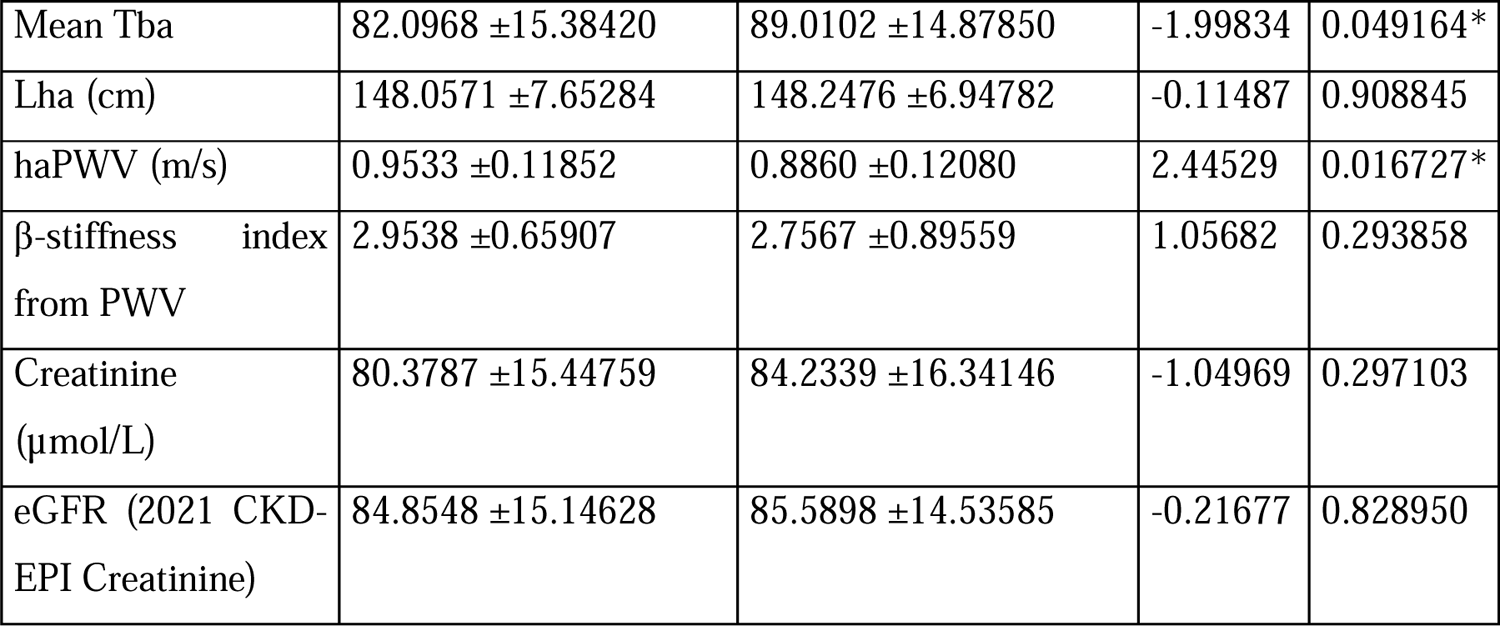
The continuous variables of the sample presented as a mean ± standard deviation (Std. div.), Student test as independent variables used. * Values statically significant difference. Abbreviations: SBP; systolic blood pressure, DBP; diastolic blood pressure, BMI, body mass index, HR; heart rate, METs; metabolic equivalent, R-CAVI; right Cardio-ankle vascular index, L-CAVI; left Cardio-ankle vascular index, RABI; right ankle-brachial index, LABI; left ankle-brachial index, SBP B; systolic blood pressure brachial, DBP B; diastolic blood pressure brachial, BP RB; blood pressure right brachial, BP RA; blood pressure right ankle, BP LA; blood pressure left ankle, BP A; blood pressure ankle, ABI; ankle-brachial index, RTb; right brachial pulse, LTb; left brachial pulse, Tb; mean brachial pulse, Tba; mean brachial-ankle pulse, Lha (cm); length heart-ankle, haPWV (m/s); heart-ankle pulse wave velocity.

A supplementary file was attached to demonstrate all the statistically significant differences between continuous variables using the binary categorical variables as a classifier. ((Suppl. 1)

### The diagnostic accuracy of the bicycle ergometry

We examined the diagnostic accuracy of a standard exercise test on a bicycle ergometer. In the ROC analysis, where the predictor was the result of a sample with the results of the physical exertion “Reaction_type” = ‘Positive’, and the target variable was Myocardial_perfusion_defect_after_stress_ATP, the following results were obtained. (*Table 3*)

**Table 3:**
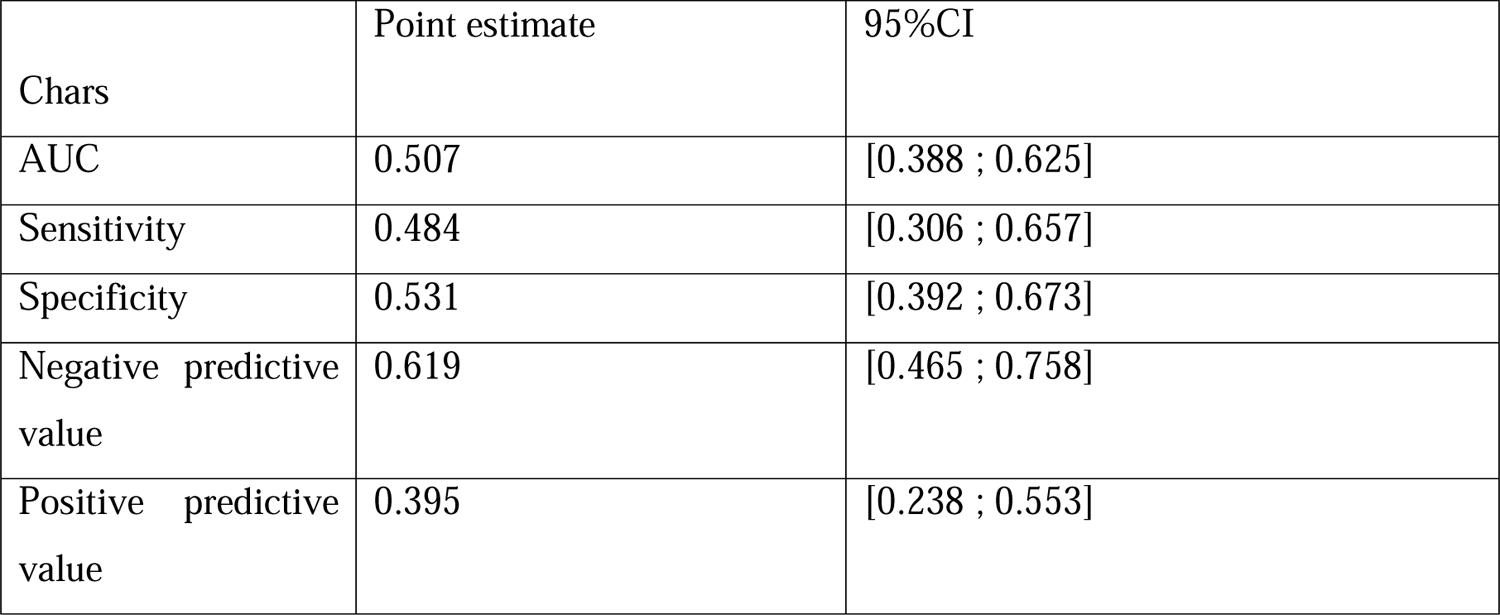
The quality of the bicycle ergometry appeared quite low in our cohort.

### Exhaled breath biomarkers diagnostic accuracy

The following predictors were selected for VOCs, the top 10 based on the median feature importances are presented below. (*Table 4*)

**Table 4:**
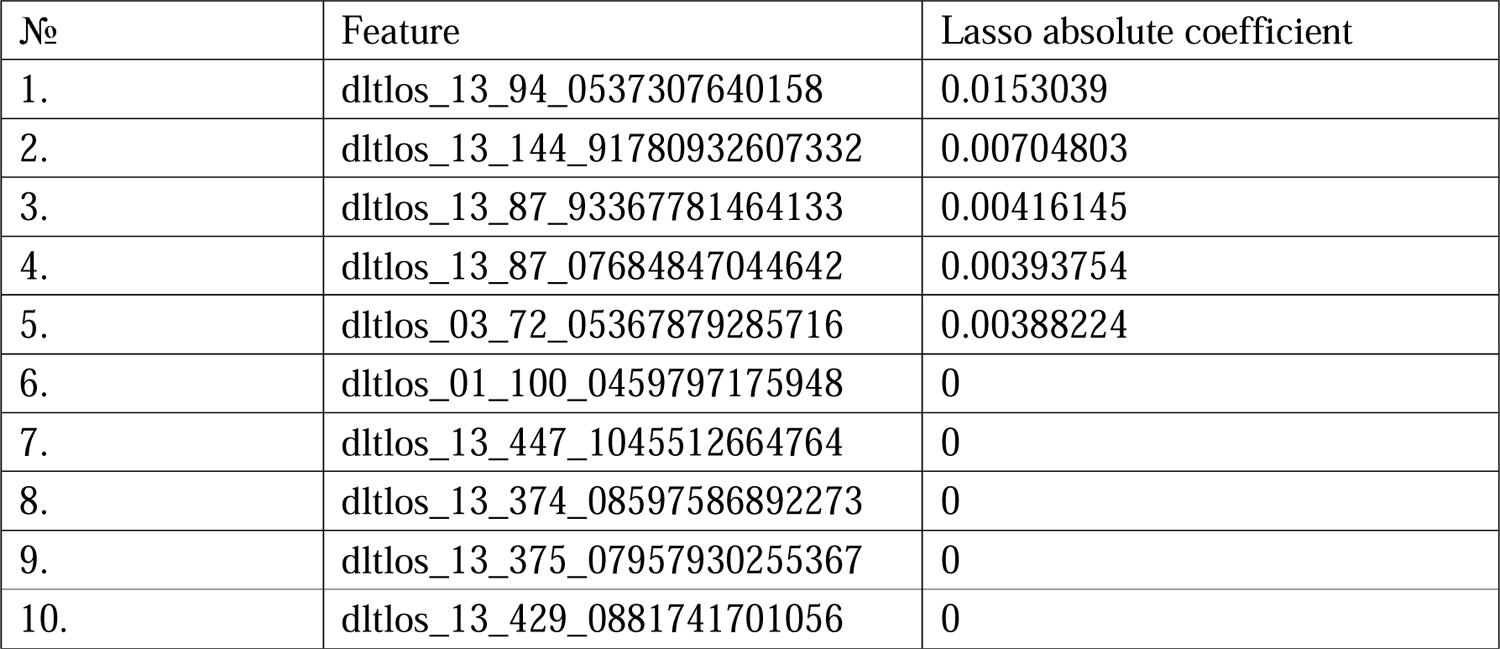
The 10 most statically significant features according to the build model represented in the table.

The model was then rebuilt as follows. The top 5 predictors from *Table 9* were with the most mathematically importance according to the built model taken and included in the new LASSO regression model.

Then the leave-one-out cross-validation procedure was performed, which allowed us to obtain approximate estimates of sensitivity, specificity, positive and negative prognostic value. At each iteration of leave-one-out cross-validation, the quantitative predictors were normalized. The quality of the classification is shown in the table below. (*Table 5*)

**Table 5:**
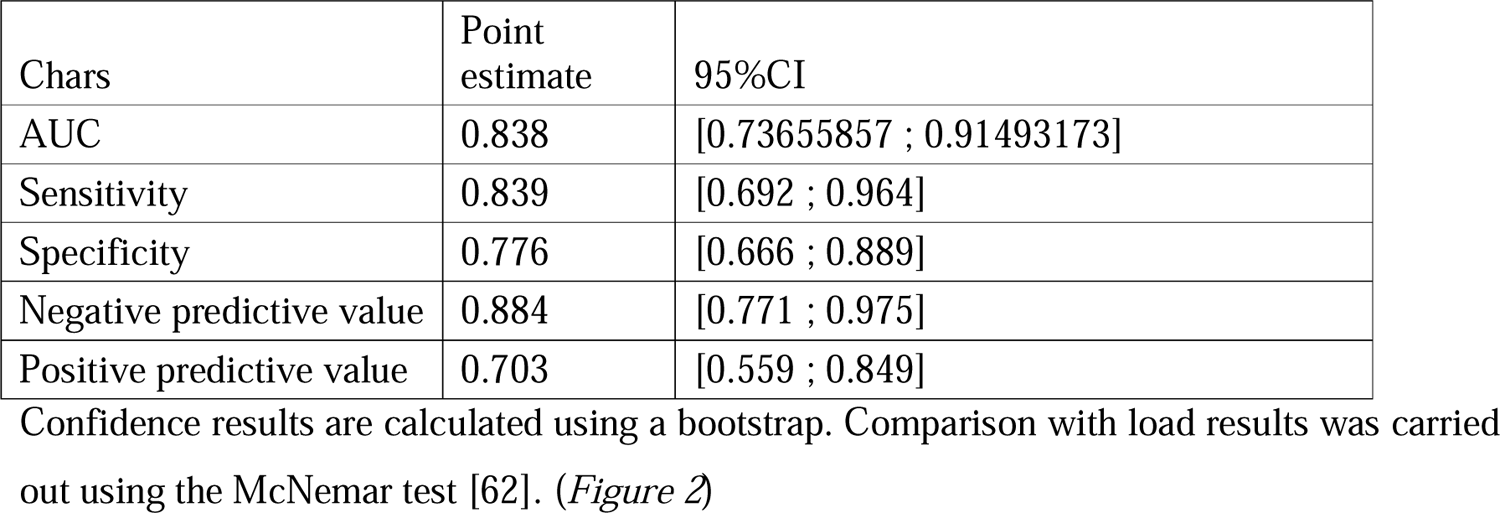
The quality of the exhaled breath biomarkers in the diagnosis of ischemic heart disease.

Confidence results are calculated using a bootstrap. Comparison with load results was carried out using the McNemar test [62]. (Figure 2)

**Figure 2:**
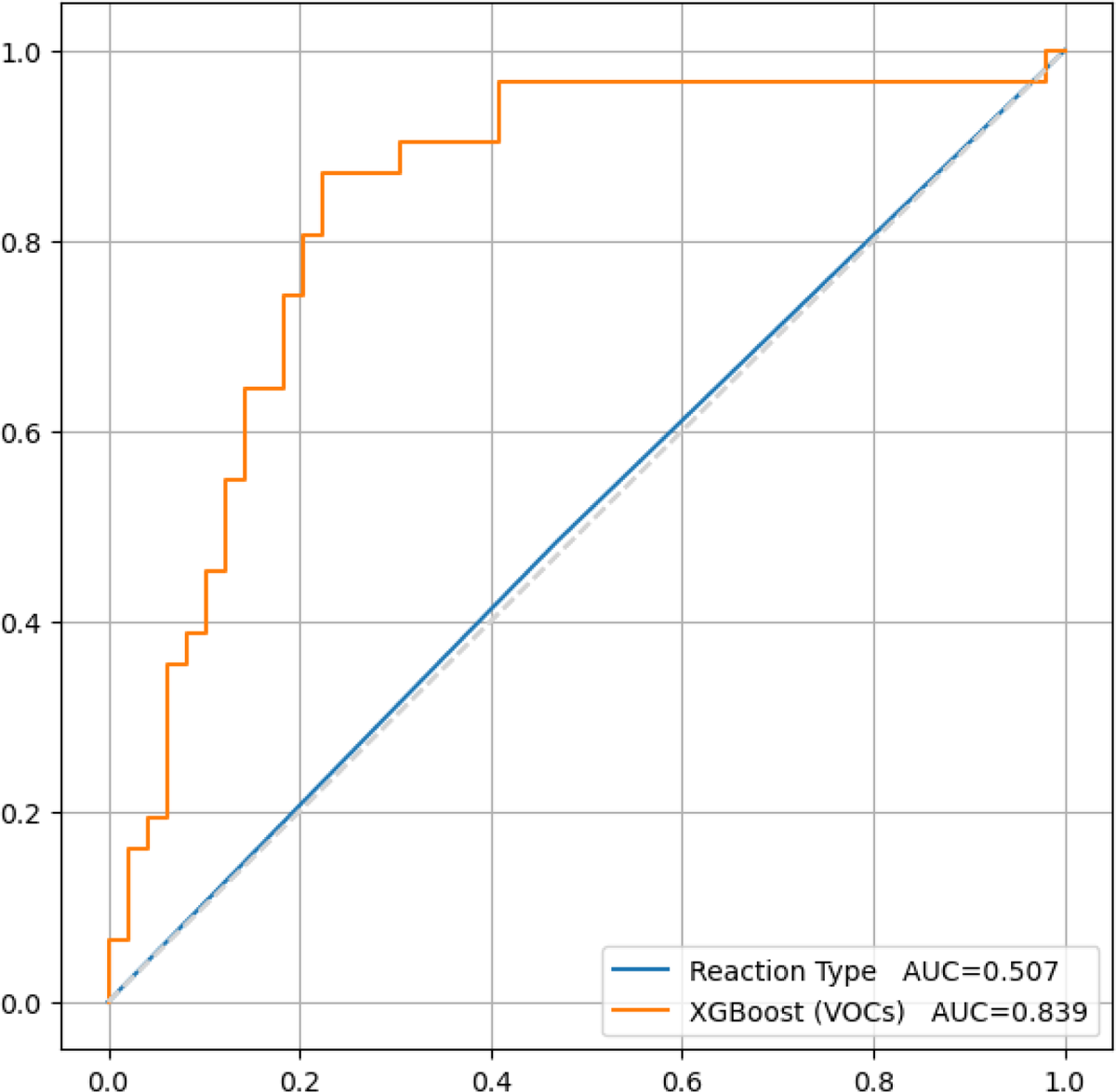
There is statistically significant difference between the results of the diagnostic accuracy of the load test (50.7 %) and the built model (83.9%), based on our study results. Obviously, the model has better predictive properties, P value =0.003.

**Figure 3:**
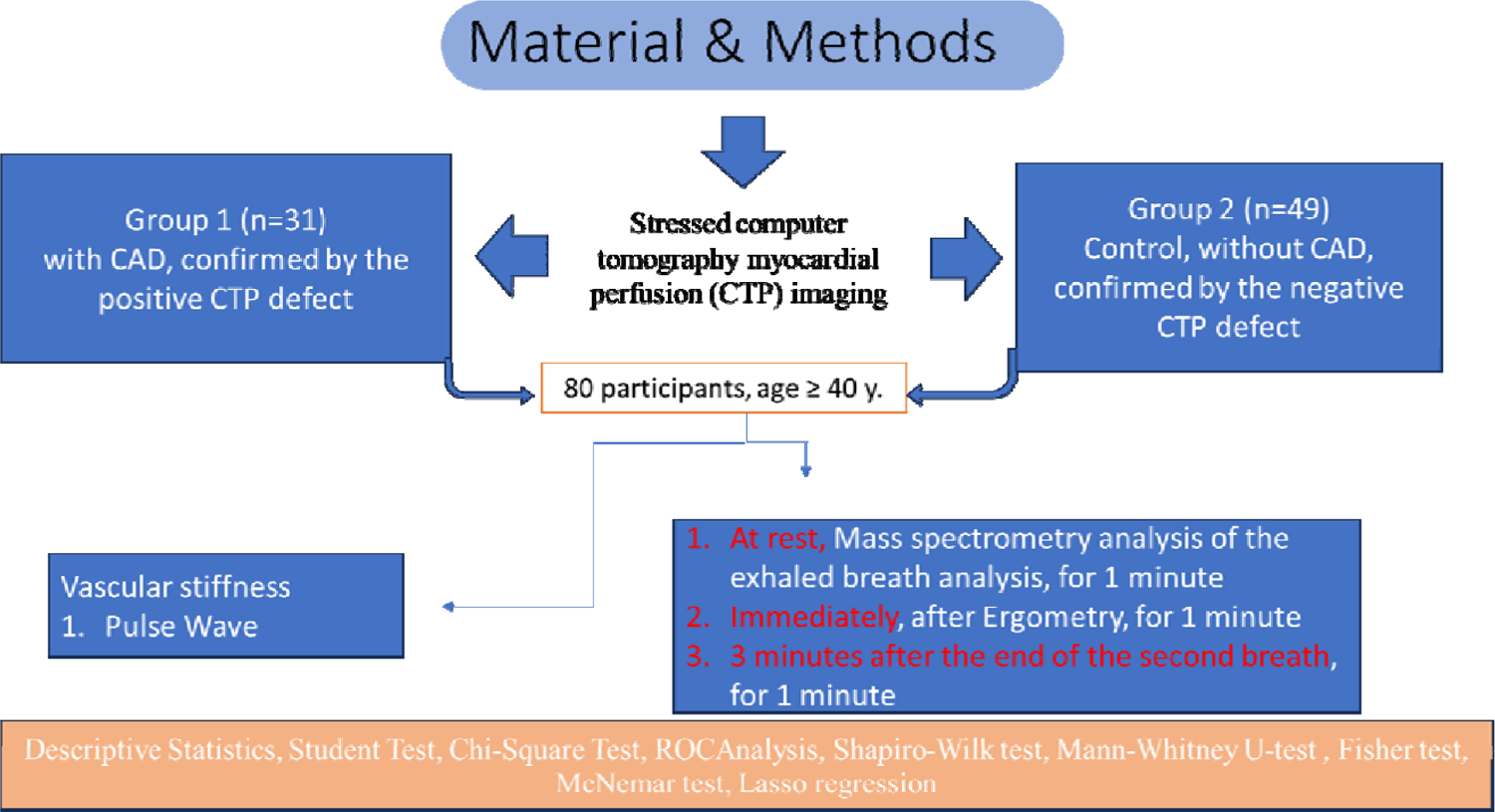
Diagrammatic presentation of the study. The patients exhale in mass spectrometer (PTR TOF-1000 (IONICON PTR-TOF-MS - Trace VOC Analyzer). Subsequently, participants pass exercise bicycle ergometry (on SCHILLER device; Bruce protocol or modified Bruce protocol). Then breath immediately after the end of the test (second breath), and after three minutes from the end of the second breath, the participants again exhale in the same mass spectrometer using single time tube.

**Figure 4:**
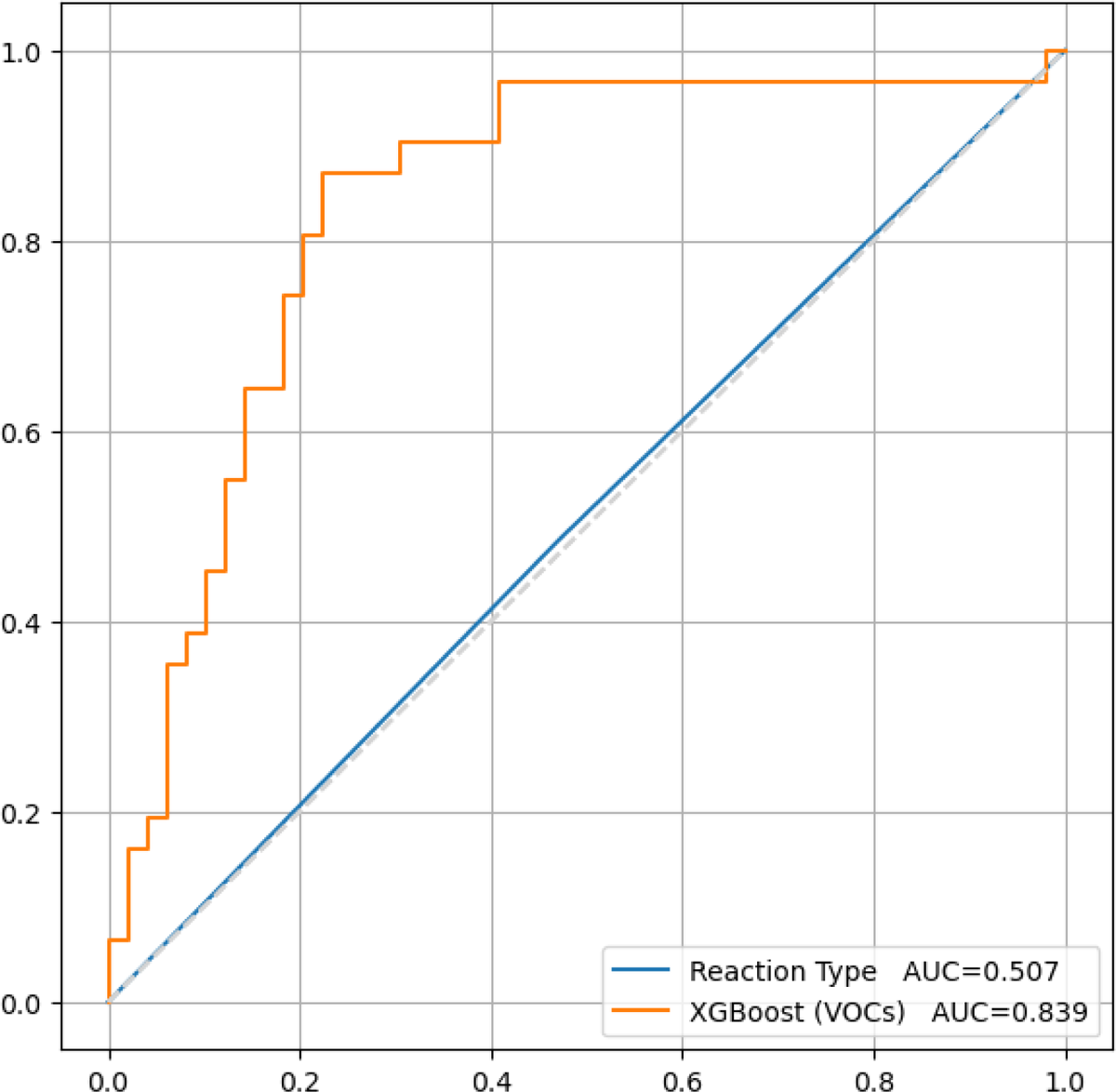
There is statistically significant difference between the results of the diagnostic accuracy of the load test (50.7 %) and the built model (83.9%), based on our study results. Obviously, the model has better predictive properties, P value =0.003.

### The found m/z annotation

The presented m/z (mass/charge) ratio has been translated into chemical formula (if exist in the literature or library of the IONICON PTR-TOF-MS-1000 device). Data preprocessing and the annotation of molecular formulae are presented in the below table. (*Table 6)*

**Table 6:**
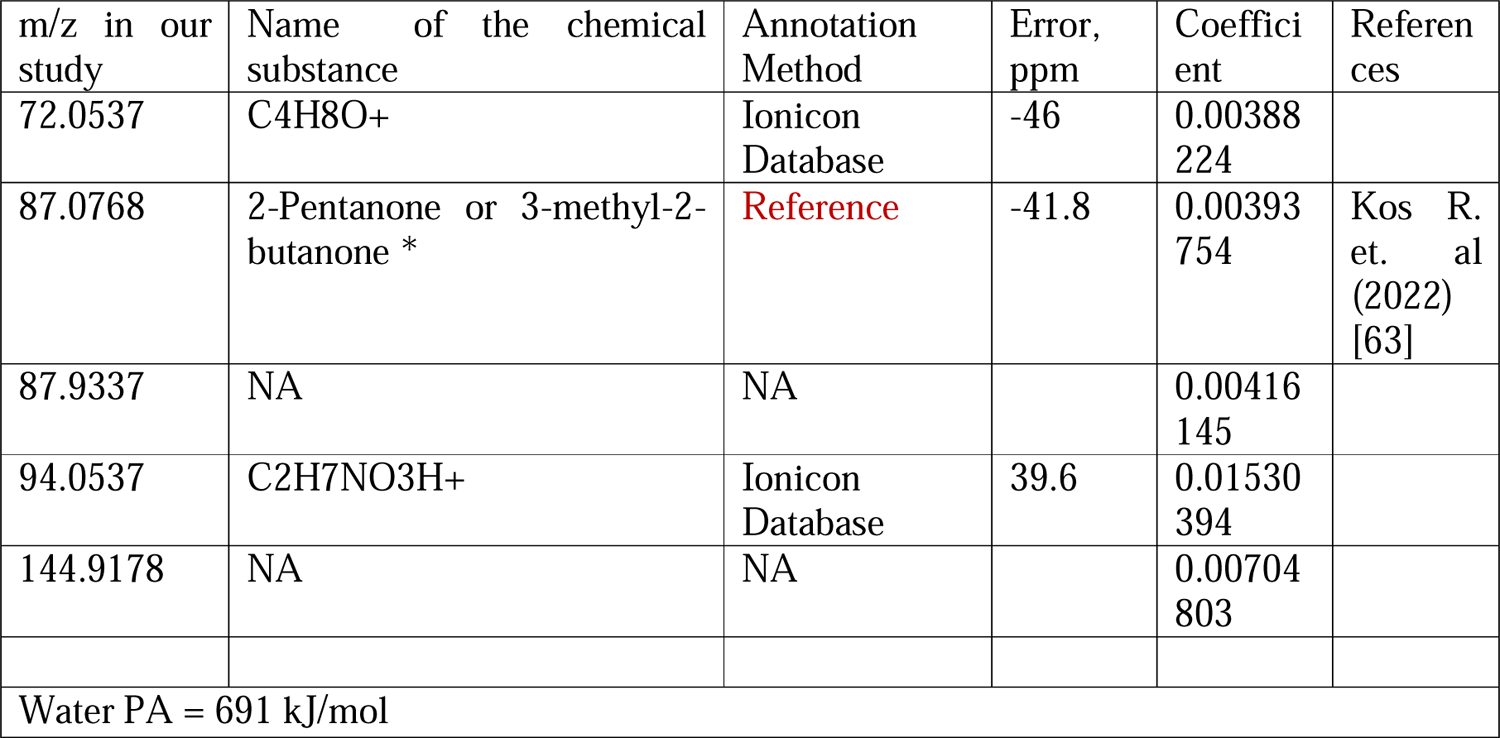
Comparative presentation of the most significantly VOCs in the exhaled breath analysis of individuals with ischemic heart disease and without ischemic heart disease. The presented m/z (mass/charge) ratios in our study are close to m/z ratios published in the literature or from the chemical substances’ library provided by the manufacturer of the IONICON PTR-TOF-MS device. In case that the m/z ratio does not have a known chemical name, represented as a chemical formula. After filtering the list of the m/z ratio from the artifacts and duplicates values, out of 10 top m/z ratios remained 5 m/z ratios. * Can be two chemical substances according to the found m/z.

**Table 7:**
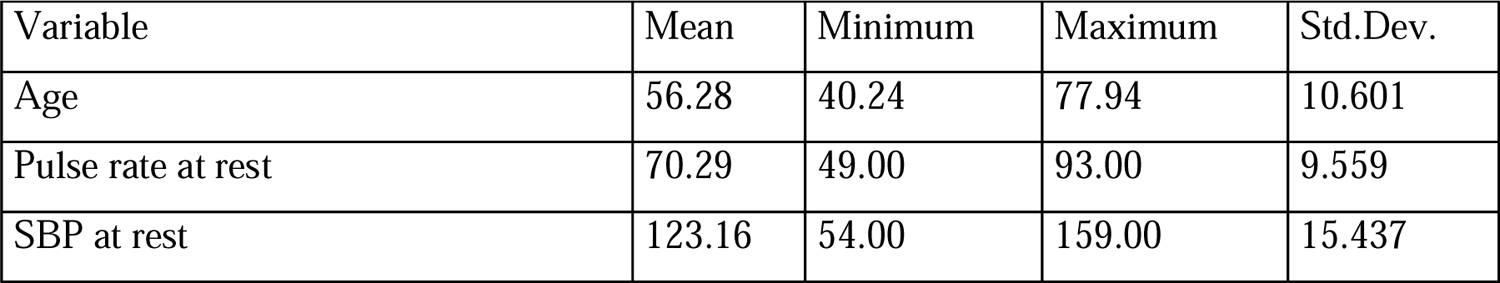
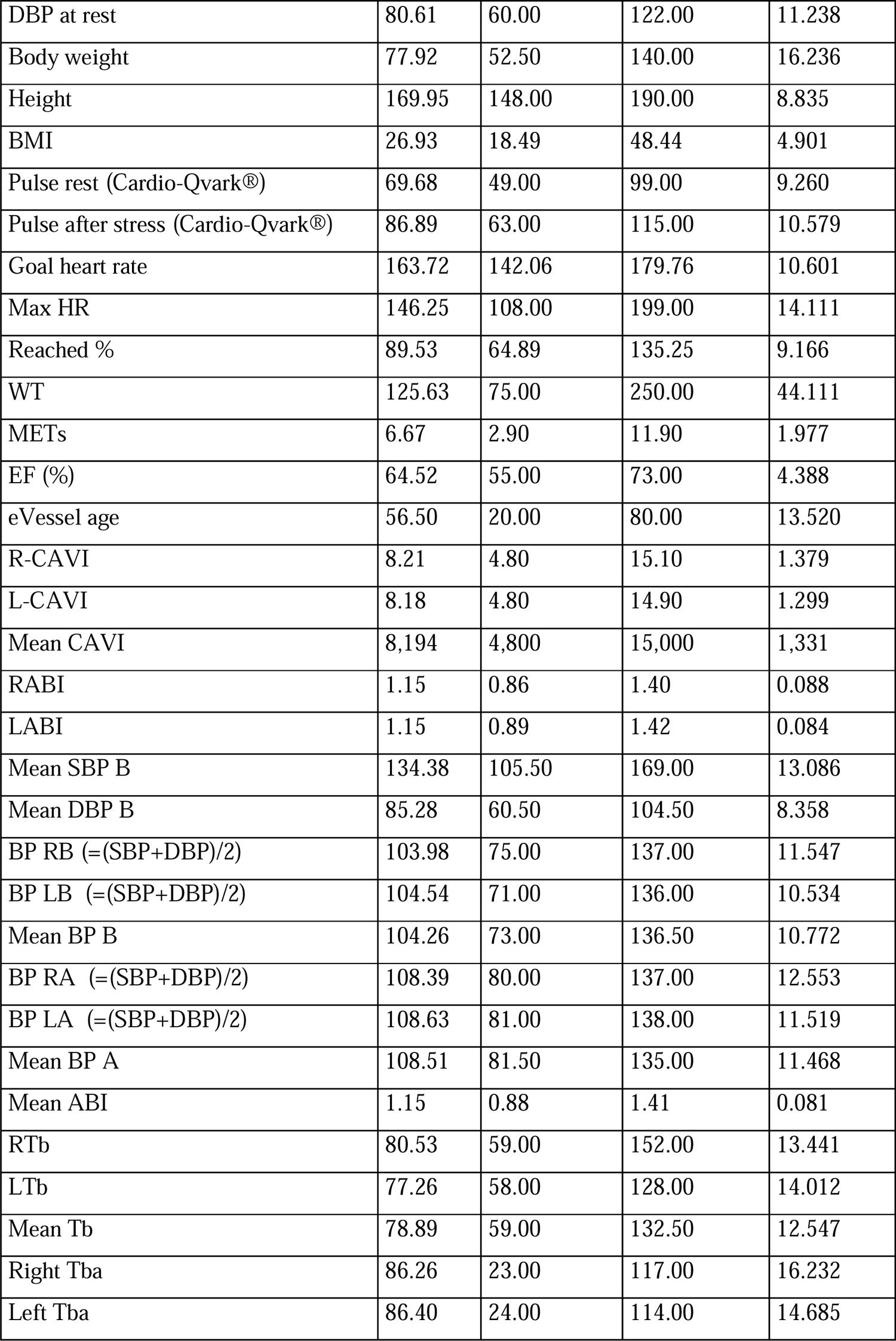
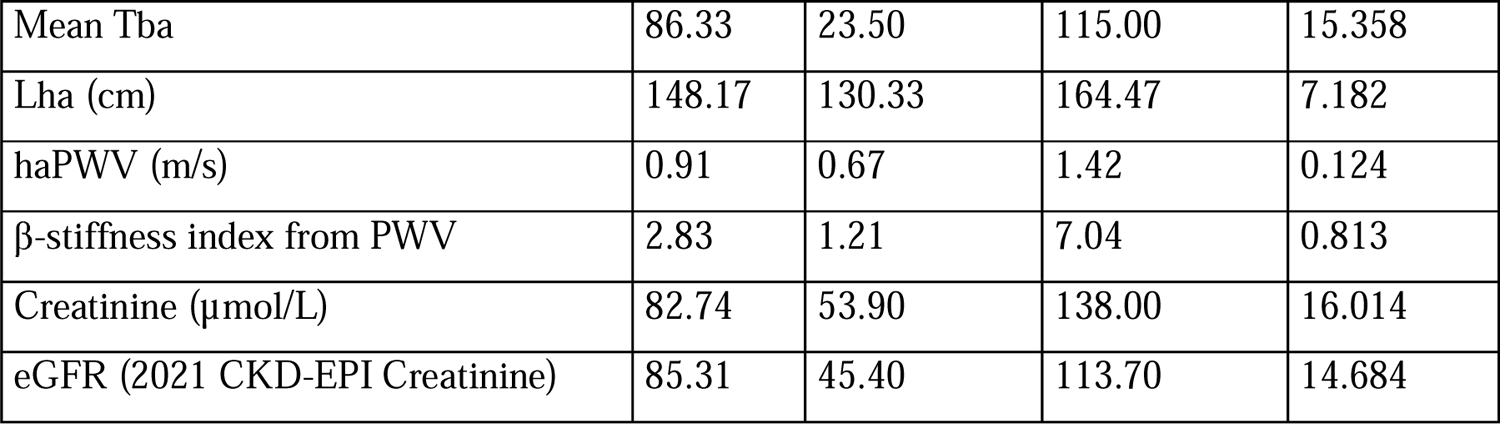
The features (Descriptive Statistics) of the continues variables of the whole sample represented in the table.

| Variable | Mean | Minimum | Maximum | Std.Dev. |
| --- | --- | --- | --- | --- |
| Age | 56.28 | 40.24 | 77.94 | 10.601 |
| Pulse rate at rest | 70.29 | 49.00 | 93.00 | 9.559 |
| SBP at rest | 123.16 | 54.00 | 159.00 | 15.437 |
| DBP at rest | 80.61 | 60.00 | 122.00 | 11.238 |
| Body weight | 77.92 | 52.50 | 140.00 | 16.236 |
| Height | 169.95 | 148.00 | 190.00 | 8.835 |
| BMI | 26.93 | 18.49 | 48.44 | 4.901 |
| Pulse rest (Cardio-Qvark®) | 69.68 | 49.00 | 99.00 | 9.260 |
| Pulse after stress (Cardio-Qvark®) | 86.89 | 63.00 | 115.00 | 10.579 |
| Goal heart rate | 163.72 | 142.06 | 179.76 | 10.601 |
| Max HR | 146.25 | 108.00 | 199.00 | 14.111 |
| Reached % | 89.53 | 64.89 | 135.25 | 9.166 |
| WT | 125.63 | 75.00 | 250.00 | 44.111 |
| METs | 6.67 | 2.90 | 11.90 | 1.977 |
| EF (%) | 64.52 | 55.00 | 73.00 | 4.388 |
| eVessel age | 56.50 | 20.00 | 80.00 | 13.520 |
| R-CAVI | 8.21 | 4.80 | 15.10 | 1.379 |
| L-CAVI | 8.18 | 4.80 | 14.90 | 1.299 |
| Mean CAVI | 8,194 | 4,800 | 15,000 | 1,331 |
| RABI | 1.15 | 0.86 | 1.40 | 0.088 |
| LABI | 1.15 | 0.89 | 1.42 | 0.084 |
| Mean SBP B | 134.38 | 105.50 | 169.00 | 13.086 |
| Mean DBP B | 85.28 | 60.50 | 104.50 | 8.358 |
| BP RB $(=(SBP+DBP)/2)$ | 103.98 | 75.00 | 137.00 | 11.547 |
| BP LB $(=(SBP+DBP)/2)$ | 104.54 | 71.00 | 136.00 | 10.534 |
| Mean BP B | 104.26 | 73.00 | 136.50 | 10.772 |
| BP RA $(=(SBP+DBP)/2)$ | 108.39 | 80.00 | 137.00 | 12.553 |
| BP LA $(=(SBP+DBP)/2)$ | 108.63 | 81.00 | 138.00 | 11.519 |
| Mean BP A | 108.51 | 81.50 | 135.00 | 11.468 |
| Mean ABI | 1.15 | 0.88 | 1.41 | 0.081 |
| RTb | 80.53 | 59.00 | 152.00 | 13.441 |
| LTb | 77.26 | 58.00 | 128.00 | 14.012 |
| Mean Tb | 78.89 | 59.00 | 132.50 | 12.547 |
| Right Tba | 86.26 | 23.00 | 117.00 | 16.232 |
| Left Tba | 86.40 | 24.00 | 114.00 | 14.685 |
| Mean Tba | 86.33 | 23.50 | 115.00 | 15.358 |
| Lha (cm) | 148.17 | 130.33 | 164.47 | 7.182 |
| haPWV (m/s) | 0.91 | 0.67 | 1.42 | 0.124 |
| β-stiffness index from PWV | 2.83 | 1.21 | 7.04 | 0.813 |
| Creatinine ($\mu\text{mol/L}$) | 82.74 | 53.90 | 138.00 | 16.014 |
| eGFR (2021 CKD-EPI Creatinine) | 85.31 | 45.40 | 113.70 | 14.684 |

**Table 8A:**
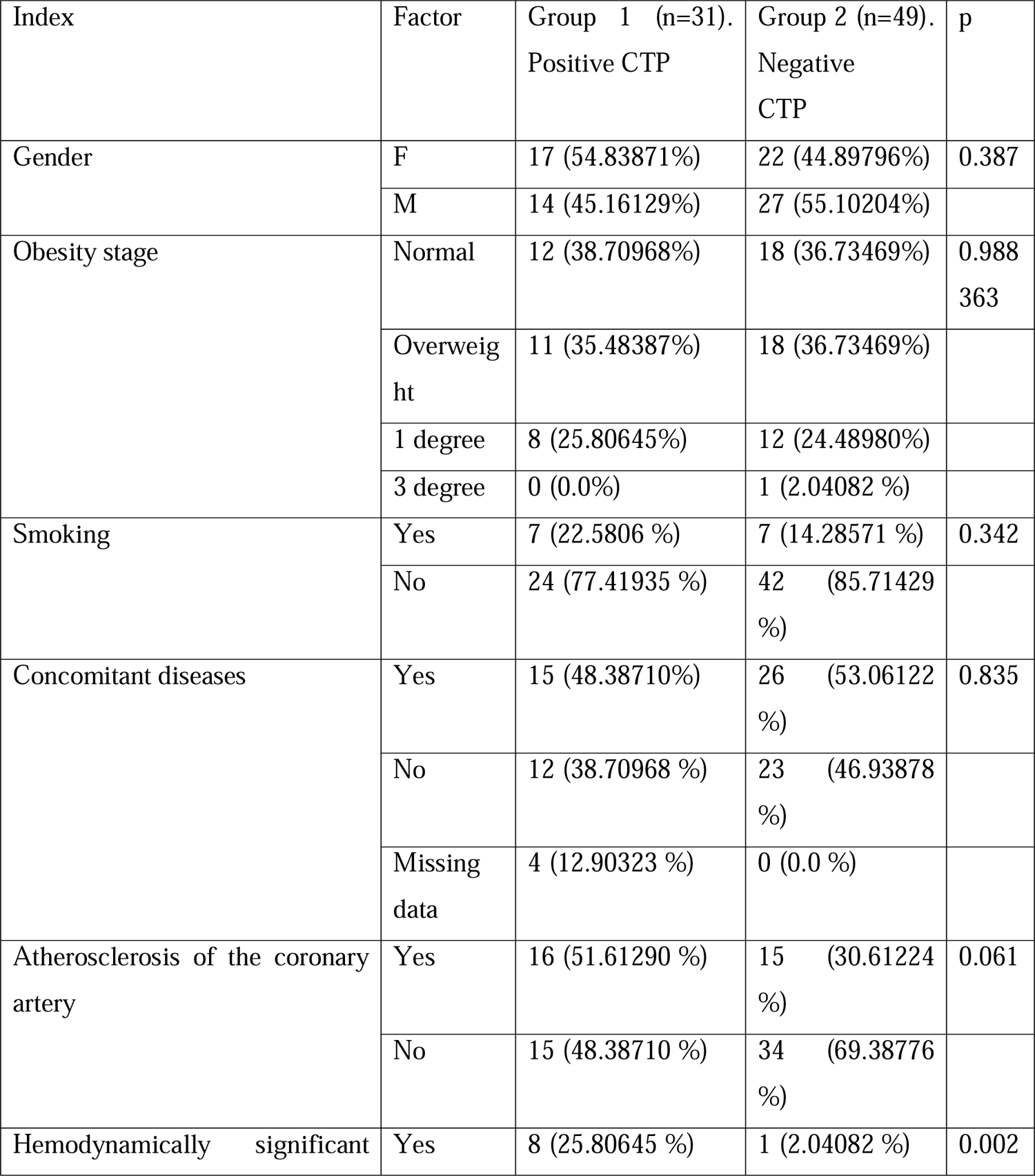
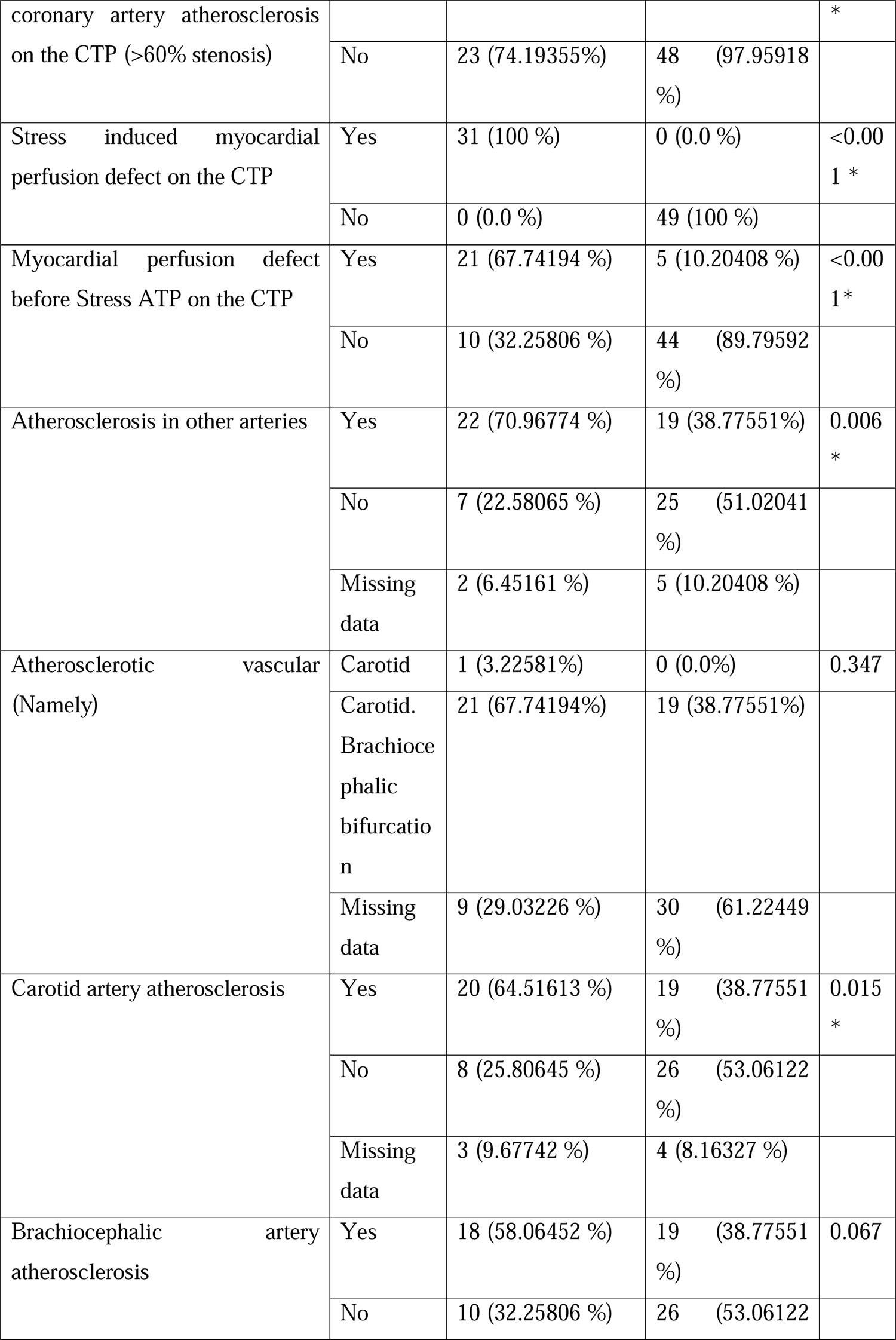
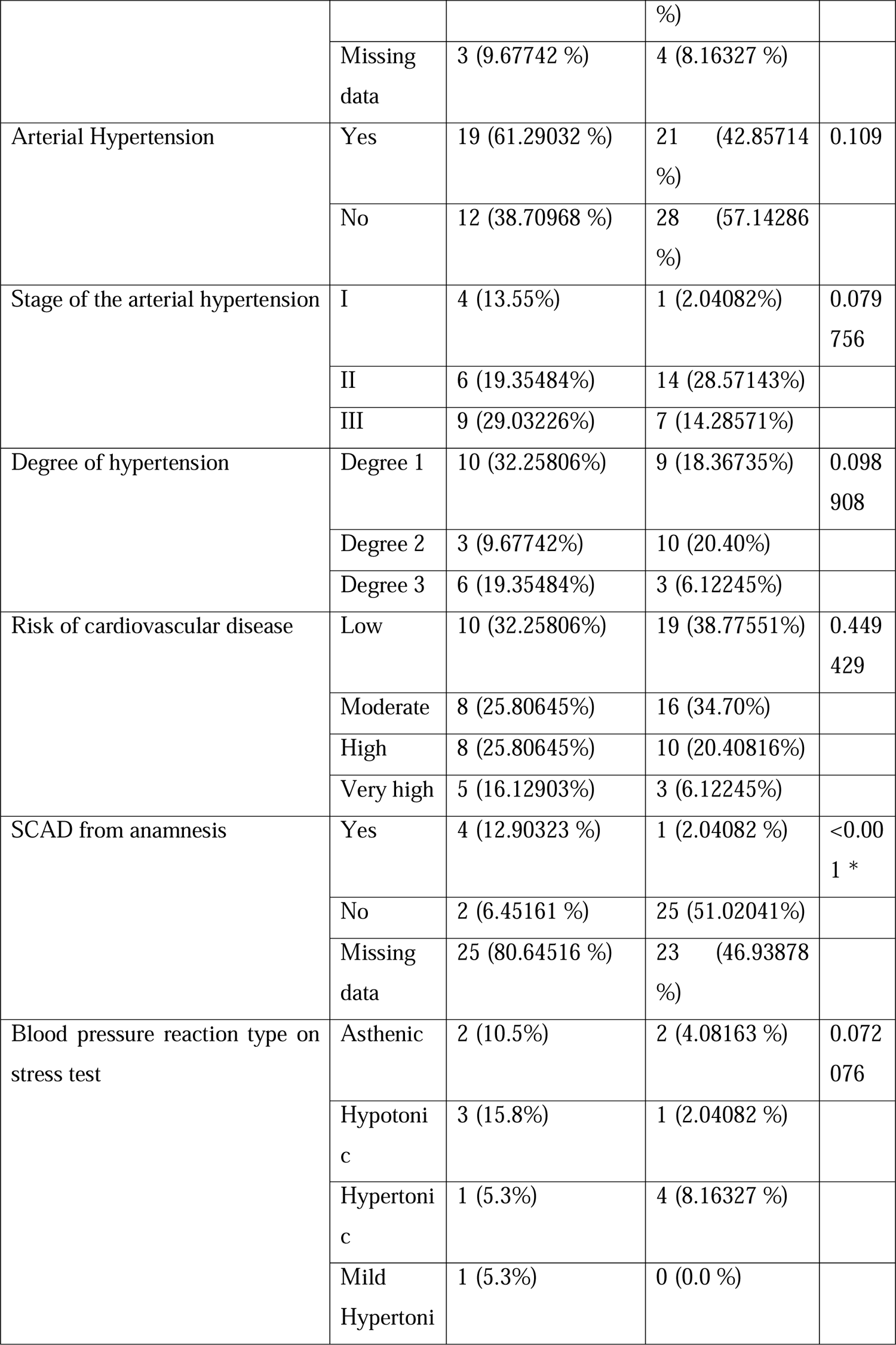
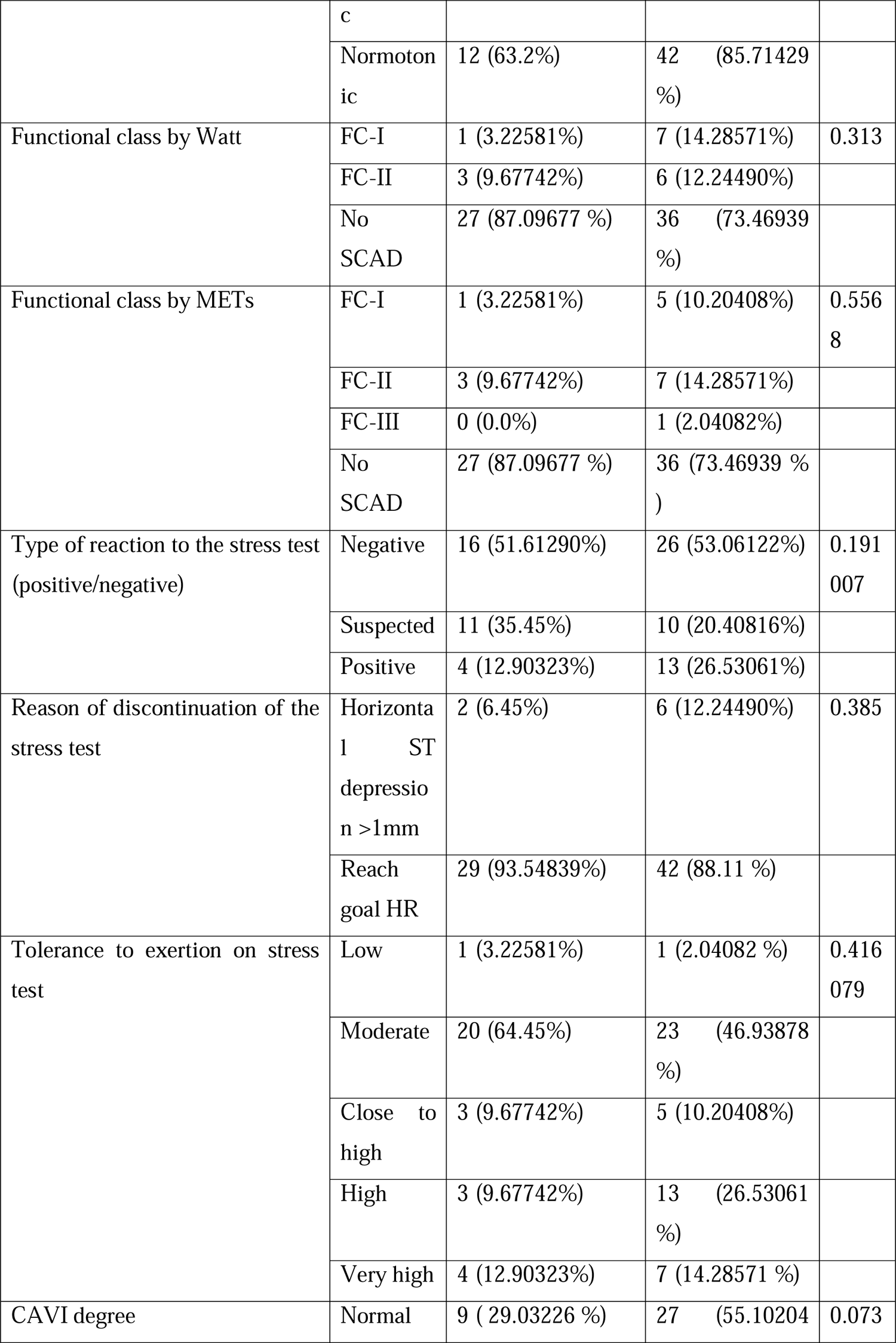
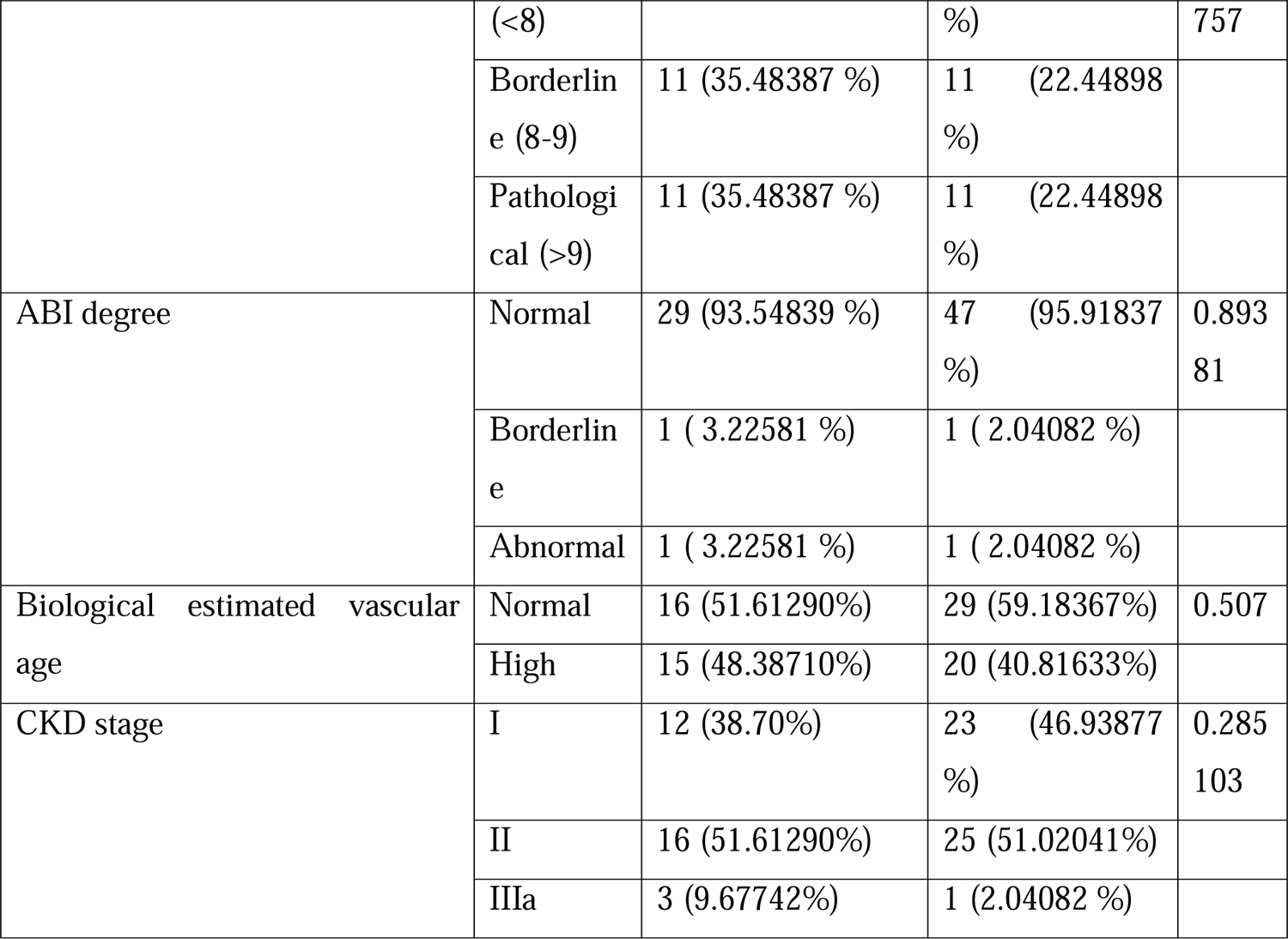
Categorical variables presented in absolute and relative values of the study for true incidence of the stated factor. X^2^ test used as a comparative test. * Values statically significant difference. Abbreviations: METs; metabolic equivalent. CPT; stress myocardial perfusion computer tomography imaging.

**Table 9:**
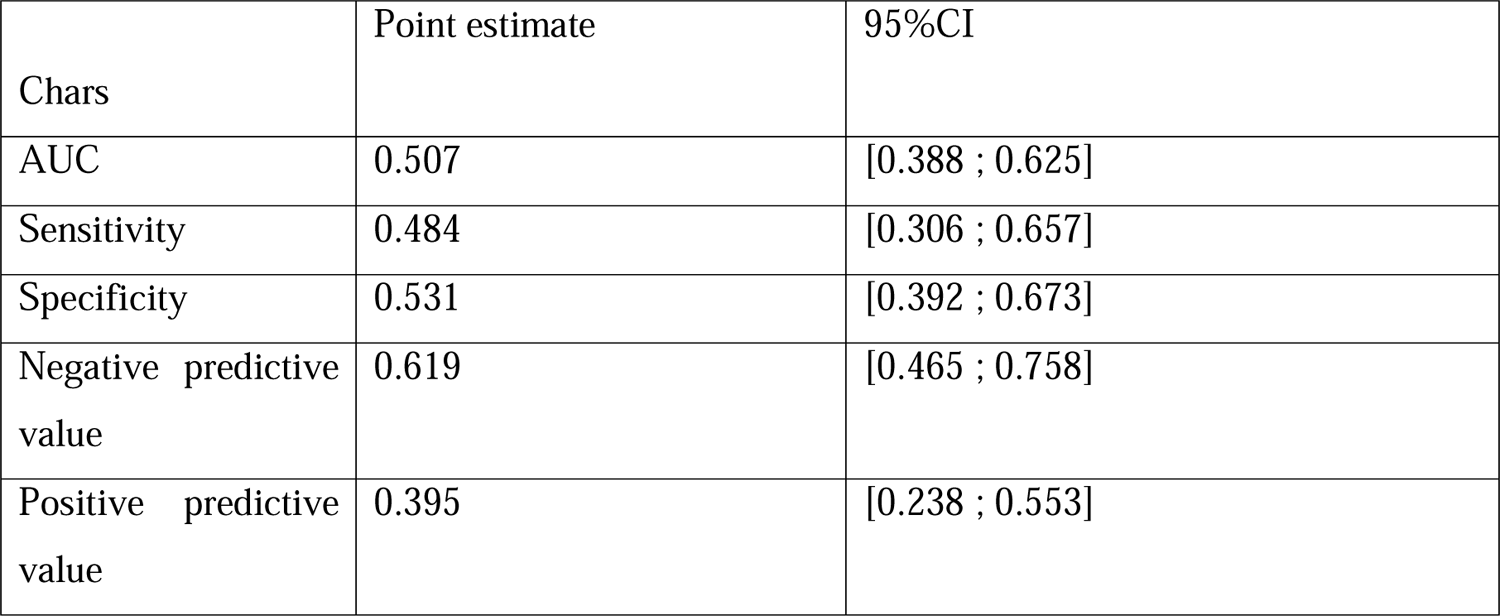
The quality of the bicycle ergometry appeared quite low in our cohort.

**Table 10:**
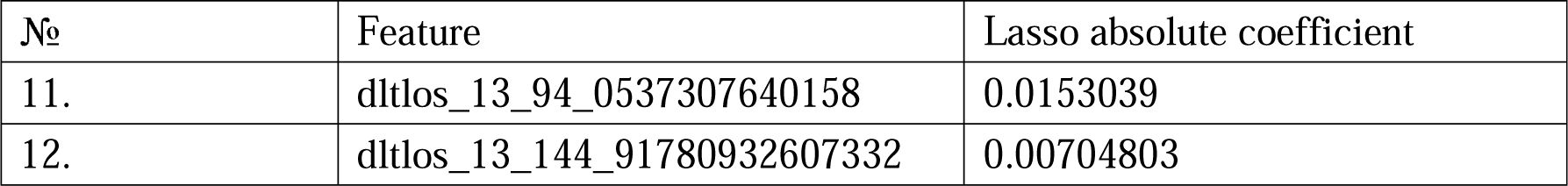
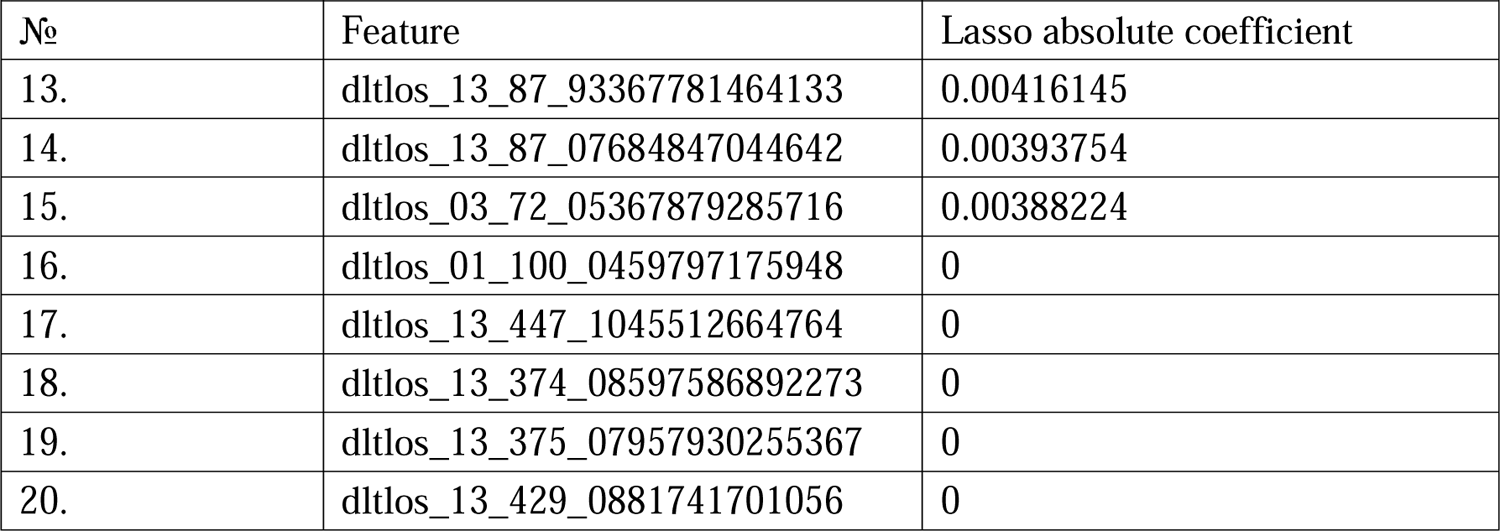
The 10 most statically significant features according to the build model represented in the table.

**Table 11:**
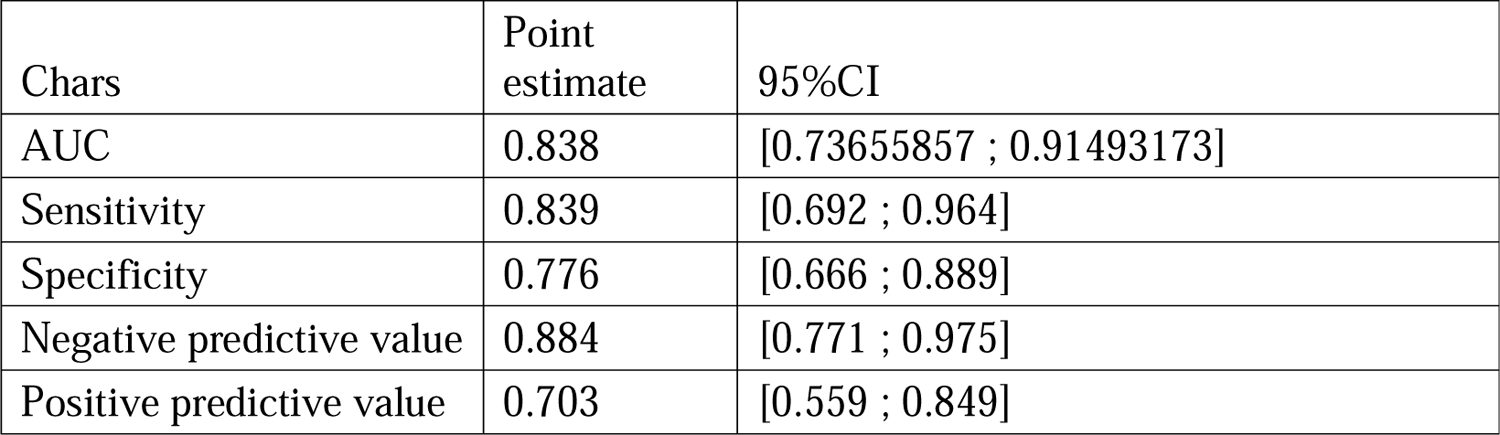
The quality of the exhaled breath biomarkers in the diagnosis of ischemic heart disease.

**Table 12:**
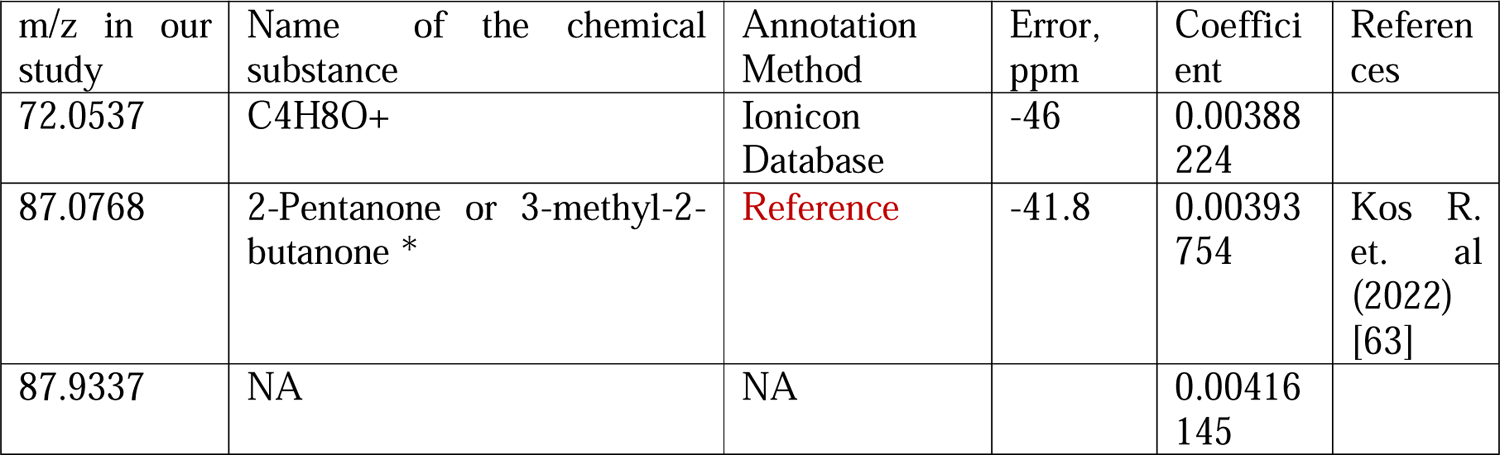
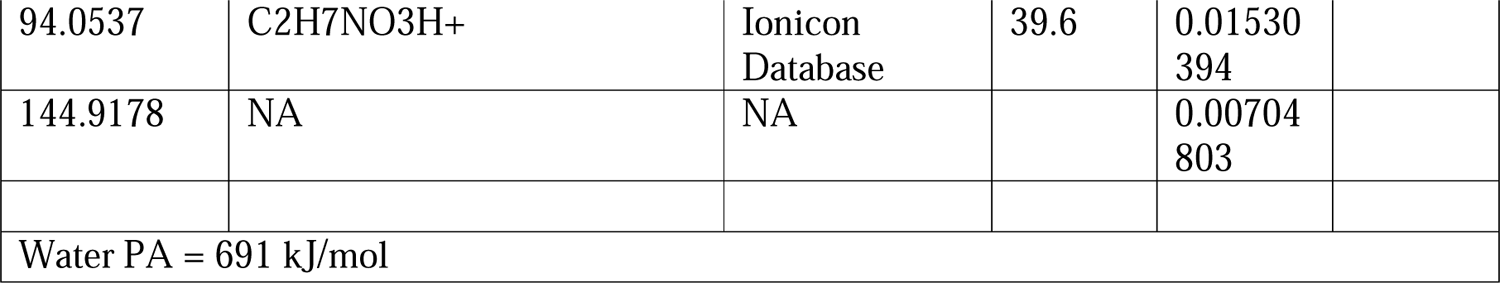
Comparative presentation of the most significantly VOCs in the exhaled breath analysis of individuals with ischemic heart disease and without ischemic heart disease. The presented m/z (mass/charge) ratios in our study are close to m/z ratios published in the literature or from the chemical substances’ library provided by the manufacturer of the IONICON PTR-TOF-MS device. In case that the m/z ratio does not have a known chemical name, represented as a chemical formula. After filtering the list of the m/z ratio from the artifacts and duplicates values, out of 10 top m/z ratios remained 5 m/z ratios. * Can be two chemical substances according to the found m/z.

PTR-TOF-MS analysis was able to detect 53 features with statically significant difference in exhaled breath from patients with ischemic heart disease and healthy controls. The m/z ratios with statistically significant difference presented in *supplementary file 1*.

An attempt is made to estimate the correlations of delts with each other. The analysis was performed between each pair of delts according to Spearman correlation test. In our study, there are a small number of patients (n=80), but a significant number of predictor variables. Routine Spearman correlation analysis revealed a huge number of correlations, while it is impossible to understand how predictors behave together. To assess the endpoint’s relationship with predictors, we used LASSO to identify the main predictors and reduce multicollinearity.

## Discussion

In the light of the presented results. the VOCs concentrations differences are statically different between the two groups (Group 1 with positive CPT/Group 2 with negative CPT). Especially, when comparing the concentration of the VOCs before and after performing the physical exertion test (bicycle ergometry), presented as a deltlos. Suggesting that changes in the concentration of the VOCs was associated with agitation of the ischemic heart disease and not for other reasons. Moreover, atherosclerosis of the other arteries, brachiocephalic or carotid, has no statistically significant difference between the groups, which confirms that the change in the concentrations of the VOCs was due to the worsening of the ischemic heart disease in terms of perturbance of the myocardial nourishment.

The changes in the concentration of the stated chemical substances statistically significant changed between the third and the second/first breath which indicates that these substance associated with the worsening the myocardial nourishment.

Fluctuation in the concentrations of VOCs are of clinical importance in the context of improvement of the diagnostic accuracy of ischemic heart disease in the clinical settings. This clinical importance is presented by the dramatical enhancement of the diagnosis of IHD in combination of physical exertion test (bicycle ergometry) and exhaled breath analysis.

The presented chemical substance is represented as a ratio to charge. Accordingly, if the mass/charge is known, we present it as a name of the chemical substance. In case the mass/charge is unknown, we leave ratio and write the chemical formula of this chemical substance.

The pathomorphological changes in ischemic heart disease represented by the metabolic acidosis of the ischemic myocardiocytes and further elaboration of the pathological cardiac metabolic changes in coronary circulation. The ischemic myocardiocytes suffers from pathophysiological changes in terms of the intercellular metabolism, which is interrupted by the disturbance in the regulation mechanisms of the intracellular homeostasis. These physiopathological changes represented by elevation of the biomarkers of the oxidative stress and the ischemia reperfusion injury.

To improve cardiometabolism, exogenous application of activators (glycolysis), activation of Sirt1 or 3 (activation of autophagy; by NAD+ administration: deacetylation), ketone oxidation, activation of the pyruvate dehydrogenase complex (glucose oxidation), and activation of the hexosamine biosynthesis pathway (O-GlcNAcylation; administration of glucosamine / glucose) pose cardio-therapeutic effects [64,65]. On the contrary, inhibition of mitochondrial oxygen consumption, malate-aspartate shuttle, mitochondrial succinate metabolism (malonate), fatty acid oxidation (CD36 inhibitors, malonyl-CoA decarboxylase inhibitors) and or inhibiting destabilization of FOF1-ATPase dimers or maintaining the association of hexokinase II or creatine kinase with mitochondria to protect the cristae structure of the mitochondria [64].

The potential source of these VOCs in the exhaled breath analysis of patients with ischemic heart disease can be assessed based on the pathomorphological changes in the ischemic heart tissue. The found VOCs include C4H8O+, 2-Pentanone or 3-methyl-2-butanone, m/z 87.9337, C2H7NO3H+, and m/z 144.9178.

## Conclusion

Exhaled breath analysis can improve the diagnostic accuracy of the bicycle ergometry in the diagnosis of IHD. Patients with discrepancy between supply and demand of heart muscle tissue with blood experience elevation in VOCs include C4H8O+, 2-Pentanone or 3-methyl-2-butanone, m/z 87.9337, C2H7NO3H+, and m/z 144.9178. Further investigation is required to reveal the full pilot of the exhaled breath analysis VOCs in patients with IHD.

## List of abbreviation

CVD: cardiovascular disease

CTP: stress computed tomography myocardial perfusion imaging

VOCs: volatile organic compounds.

## Decelerations

1. Ethics approval and consent to participate: the study approved by the Sechenov University, Russia, from “Ethics Committee Requirement № 19-23 from 26.10.2023”. A written consent is taken from the study participants.
2. Consent for publication: applicable on reasonable request
3. Availability of data and materials: applicable on reasonable request
4. Competing interests: The authors declare that they have no competing interests regarding publication.
5. Funding’s: The work financed by the Ministry of Science and Higher Education of the Russian Federation within the framework of state support for the creation and development of World-Class Research Center ‘Digital biodesign and personalized healthcare’ № 075-15-2022-304.
6. Authors’ contributions: MB is the writer, researcher, collected and analyzed data, interpreted the results. and revised the final version of the paper, AS chemical analyze of the exhaled breath, AS biostatistical analysis of the sample, PCh, DG, NVG, EF revised the paper, and PhK revised the final version of the manuscript. All authors have read and approved the manuscript.

## Data Availability

All data produced in the present study are available upon reasonable request to the authors

## Acknowledgments

not applicable

## Authors’ information

**Basheer Abdullah Marzoog**, World-Class Research Center «Digital Biodesign and Personalized Healthcare», I.M. Sechenov First Moscow State Medical University (Sechenov University), 119991 Moscow, Russia; postal address: Russia. Moscow, 8-2 Trubetskaya street, 119991, (, +79969602820). ORCID: 0000-0001-5507-2413. Scopus ID: 57486338800. **Peter Chomakhidze**, World-Class Research Center «Digital Biodesign and Personalized Healthcare», I.M. Sechenov First Moscow State Medical University (Sechenov University), 119991 Moscow, Russia; postal address: Russia. Moscow, 8-2 Trubetskaya street, 119991. ORCID: 0000-0003-1485-6072. **Daria Gognieva**, World-Class Research Center «Digital Biodesign and Personalized Healthcare», I.M. Sechenov First Moscow State Medical University (Sechenov University), 119991 Moscow, Russia; postal address: Russia. Moscow, 8-2 Trubetskaya street, 119991. ORCID: 0000-0002-0451-2009. **Nina Vladimirovna Gagarina,** University clinical Hospital number 1, Radiology department, I.M. Sechenov First Moscow State Medical University (Sechenov University), 119991 Moscow, Russia; postal address: Russia, Moscow, 8-2 Trubetskaya street, 119991. Scopus ID: 6508312251. **Artemiy Silantyev,** World-Class Research Center «Digital Biodesign and Personalized Healthcare», I.M. Sechenov First Moscow State Medical University (Sechenov University), 119991 Moscow, Russia; postal address: Russia. Moscow, 8-2 Trubetskaya street, 119991. ORCID: 0000-0002-0451-2009. **Alexander Suvorov,** World-Class Research Center «Digital Biodesign and Personalized Healthcare», I.M. Sechenov First Moscow State Medical University (Sechenov University), 119991 Moscow, Russia; postal address: Russia. Moscow, 8-2 Trubetskaya street, 119991. ORCID: 0000-0002-0451-2009. **Ekaterina Fominykha**, University clinical Hospital number 1, Radiology department, I.M. Sechenov First Moscow State Medical University (Sechenov University), 119991 Moscow, Russia; postal address: Russia, Moscow, 8-2 Trubetskaya street, 119991, ORCID: 0000-0003-0288-7656. **Philipp Kopylov,** director of the institute of the Research Center «Digital Biodesign and Personalized Healthcare», World-Class Research Center «Digital Biodesign and Personalized Healthcare», I.M. Sechenov First Moscow State Medical University (Sechenov University), 119991 Moscow, Russia; postal address: Russia. Moscow, 8-2 Trubetskaya street, 119991. ORCID: 0000-0002-4535-8685. Scopus ID: 6507736224. The paper has not been submitted elsewhere

## STANDARDS OF REPORTING

STROBE guideline has been followed.

---

**Supplementary 1:** The comparative features of the sample divided by various binary categorical variables. The represented continuous variables are all statistically significant at p<0.05.

| Variable | T-tests; Grouping: Gender (Both in primary data.stw) |  |  |  |
| --- | --- | --- | --- | --- |
|  | Group 1: F<br>Group 2: M |  |  |  |
|  | Mean<br>F | Mean<br>M | t-value | p |
| Body weight | 71.5974 | 83.9341 | -3.65280 | 0.000468 |
| Height | 163.9359 | 175.6707 | -7.93010 | 0.000000 |
| WT | 94.8718 | 154.8780 | -8.28672 | 0.000000 |
| METs | 5.8846 | 7.4195 | -3.74534 | 0.000343 |
| Lha (cm) | 143.2849 | 152.8242 | -7.93010 | 0.000000 |
| β-stiffness index from PWV | 2.6430 | 3.0138 | -2.08085 | 0.040730 |
| Creatinine ($\mu\text{mol/L}$) | 72.4862 | 92.4937 | -7.13522 | 0.000000 |

B
| Variable | T-tests; Grouping: Smoking (Both in primary data.stw) |  |  |  |
| --- | --- | --- | --- | --- |
|  | Group 1: No<br>Group 2: Yes |  |  |  |
|  | Mean<br>No | Mean<br>Yes | t-value | p |
| DBP rest | 79.4394 | 86.1429 | -2.06890 | 0.041869 |
| Body weight | 75.3121 | 90.2143 | -3.31009 | 0.001414 |
| Height | 168.9848 | 174.5000 | -2.17081 | 0.032987 |
| BMI | 26.3829 | 29.4905 | -2.20717 | 0.030242 |
| Lha (cm) | 147.3892 | 151.8725 | -2.17081 | 0.032987 |

C
| Variable | T-tests; Grouping: Concomitant disease (Both in primary data.stw) |  |  |  |
| --- | --- | --- | --- | --- |
|  | Group 1: Yes<br>Group 2: No |  |  |  |
|  | Mean<br>Yes | Mean<br>No | t-value | p |
| Age | 58.2005 | 53.0605 | 2.15911 | 0.034084 |
| BMI | 28.2638 | 25.4612 | 2.54843 | 0.012894 |
| Goal heart rate | 161.7995 | 166.9395 | -2.15911 | 0.034084 |
| WT | 115.8537 | 139.2857 | -2.35097 | 0.021390 |
| METs | 6.0683 | 7.5000 | -3.29168 | 0.001528 |
| eVessel age | 59.4146 | 52.0286 | 2.42323 | 0.017829 |
| Mean SBP B | 136.6829 | 130.7429 | 2.02434 | 0.046545 |
| BP RA<br>(=(SBP+DBP)/2) | 110.7561 | 104.0571 | 2.42353 | 0.017815 |
| BP LA (=(SBP+DBP)/2) | 111.2195 | 103.9143 | 3.02442 | 0.003423 |
| Mean BP A | 110.9878 | 103.9857 | 2.85424 | 0.005593 |
| RTb | 77.7805 | 84.6286 | -2.24257 | 0.027920 |
| Mean Tb | 76.3780 | 82.7714 | -2.24909 | 0.027482 |

D
| Variable | T-tests; Grouping: Atherosclerosis of the coronary artery (Both in primary data.stw)<br>Group 1: No<br>Group 2: Yes |  |  |  |
| --- | --- | --- | --- | --- |
|  | Mean No | Mean Yes | t-value | p |
| Age | 51.2944 | 64.1558 | -6.53443 | 0.000000 |
| Body weight | 74.1694 | 83.8484 | -2.69905 | 0.008522 |
| Goal heart rate | 168.7056 | 155.8442 | 6.53443 | 0.000000 |
| Reached % | 87.7888 | 92.2900 | -2.19067 | 0.031461 |
| eVessel age | 50.1224 | 66.5806 | -6.56896 | 0.000000 |
| R-CAVI | 7.6796 | 9.0419 | -4.89037 | 0.000005 |
| L-CAVI | 7.7122 | 8.9226 | -4.53296 | 0.000021 |
| Mean SBP B | 131.3673 | 139.1290 | -2.68406 | 0.008879 |
| BP RB $(=(SBP+DBP)/2)$ | 101.1020 | 108.5161 | -2.92887 | 0.004458 |
| BP LB $(=(SBP+DBP)/2)$ | 102.0000 | 108.5484 | -2.82609 | 0.005982 |
| Mean BP B | 101.5510 | 108.5323 | -2.95961 | 0.004078 |
| LTb | 79.7143 | 73.3871 | 2.00482 | 0.048452 |
| Right Tba | 91.5714 | 77.8710 | 4.01447 | 0.000136 |
| Left Tba | 91.1224 | 78.9355 | 3.93370 | 0.000180 |
| Mean Tba | 91.3469 | 78.4032 | 4.00722 | 0.000139 |
| haPWV (m/s) | 0.8614 | 0.9922 | -5.36049 | 0.000001 |
| β-stiffness index from PWV | 2.5866 | 3.2226 | -3.66533 | 0.000449 |
| Creatinine (μmol/L) | 79.2967 | 88.1826 | -2.49669 | 0.014644 |
| eGFR (2021 CKD-EPI Creatinine) | 88.9265 | 79.5806 | 2.90054 | 0.004838 |

E
| Variable | T-tests; Grouping: Hemodynamically significant (>60%) (Both in primary data.stw)<br>Group 1: No<br>Group 2: Yes |  |  |  |
| --- | --- | --- | --- | --- |
|  | Mean No | Mean Yes | t-value | p |
| Age | 54.6859 | 68.8402 | -4.14128 | 0.000087 |
| Goal heart rate | 165.3141 | 151.1598 | 4.14128 | 0.000087 |
| Max HR | 148.2254 | 130.6667 | 3.80483 | 0.000281 |
| EF (%) | 65.1207 | 59.5714 | 3.41368 | 0.001125 |
| eVessel age | 54.4085 | 73.0000 | -4.29401 | 0.000050 |
| R-CAVI | 7.9563 | 10.1889 | -5.30446 | 0.000001 |
| L-CAVI | 7.9634 | 9.9000 | -4.75273 | 0.000009 |
| Mean SBP B | 133.0423 | 144.8889 | -2.65461 | 0.009621 |
| Mean DBP B | 84.4014 | 92.1667 | -2.73106 | 0.007803 |
| BP RB<br>(=(SBP+DBP)/2) | 102.6620 | 114.3333 | -2.99746 | 0.003650 |
| BP LB<br>(=(SBP+DBP)/2) | 103.5070 | 112.6667 | -2.54098 | 0.013038 |
| Mean BP B | 103.0845 | 113.5000 | -2.85369 | 0.005532 |
| LTb | 78.3803 | 68.4444 | 2.04390 | 0.044339 |
| Mean Tb | 79.8944 | 71.0000 | 2.04336 | 0.044394 |
| Right Tba | 88.9718 | 64.8889 | 4.72565 | 0.000010 |
| Left Tba | 88.7606 | 67.7778 | 4.50455 | 0.000023 |
| Mean Tba | 88.8662 | 66.3333 | 4.65823 | 0.000013 |
| haPWV (m/s) | 0.8871 | 1.1089 | -6.13269 | 0.000000 |
| β-stiffness index from PWV | 2.6918 | 3.9474 | -4.97558 | 0.000004 |

F
| Variable | T-tests; Grouping: Myocardial perfusion defect before Stress ATP (Both in primary data.stw)<br>Group 1: Yes<br>Group 2: No |  |  |  |  |
| --- | --- | --- | --- | --- | --- |
|  | Mean Yes | Mean No | t-value | df | p |
| Max HR | 138.4615 | 150.0000 | -3.68868 | 78 | 0.000415 |
| Reached % | 86.4746 | 91.0055 | -2.11588 | 78 | 0.037545 |
| EF (%) | 62.7000 | 65.3333 | -2.30735 | 63 | 0.024333 |
| eVessel age | 61.8846 | 53.9074 | 2.55701 | 78 | 0.012498 |
| LABI | 1.1204 | 1.1602 | -2.03157 | 78 | 0.045603 |
| Right Tba | 80.3846 | 89.0926 | -2.30823 | 78 | 0.023637 |
| Mean Tba | 81.2115 | 88.7963 | -2.11386 | 78 | 0.037723 |

G
| Variable | T-tests; Grouping: Atherosclerosis in other arteries (Yes/No) (Both in primary data.stw)<br>Group 1: No<br>Group 2: Yes |  |  |  |
| --- | --- | --- | --- | --- |
|  | Mean No | Mean Yes | t-value | p |
| Age | 48.4591 | 61.8114 | -6.67105 | 0.000000 |
| SBP rest | 116.8750 | 126.9024 | -2.81141 | 0.006372 |
| DBP rest | 77.0313 | 82.8780 | -2.21050 | 0.030294 |
| Body weight | 71.1656 | 80.9585 | -2.71672 | 0.008276 |
| BMI | 24.4660 | 28.4944 | -3.74890 | 0.000359 |
| Goal heart rate | 171.5409 | 158.1886 | 6.67105 | 0.000000 |
| Max HR | 152.4063 | 141.3415 | 3.45324 | 0.000938 |
| WT | 139.0625 | 114.6341 | 2.35423 | 0.021332 |
| METs | 7.9406 | 5.8195 | 5.19338 | 0.000002 |
| eVessel age | 46.3125 | 64.0000 | -7.21404 | 0.000000 |
| R-CAVI | 7.5125 | 8.6780 | -3.85771 | 0.000249 |
| L-CAVI | 7.5781 | 8.5707 | -3.44625 | 0.000959 |
| Mean SBP B | 127.1406 | 139.8171 | -4.41075 | 0.000036 |
| Mean DBP B | 80.3438 | 88.1463 | -4.39502 | 0.000038 |
| BP RB<br>(=(SBP+DBP)/2) | 97.2500 | 108.5122 | -4.51995 | 0.000024 |
| BP LB<br>(=(SBP+DBP)/2) | 99.3125 | 107.9512 | -3.63760 | 0.000518 |
| Mean BP B | 98.2813 | 108.2317 | -4.21410 | 0.000073 |
| BP RA<br>(=(SBP+DBP)/2) | 101.0000 | 112.6098 | -4.38235 | 0.000040 |
| BP LA<br>(=(SBP+DBP)/2) | 101.7188 | 113.0000 | -4.69401 | 0.000013 |
| Mean BP A | 101.3594 | 112.8049 | -4.81404 | 0.000008 |
| RTb | 87.6563 | 76.0488 | 3.97243 | 0.000169 |
| LTb | 82.4063 | 74.5366 | 2.40142 | 0.018952 |
| Mean Tb | 85.0312 | 75.2927 | 3.48807 | 0.000839 |
| Right Tba | 94.5938 | 79.2439 | 4.35025 | 0.000045 |
| Left Tba | 93.8750 | 80.2927 | 4.22749 | 0.000069 |
| Mean Tba | 94.2344 | 79.7683 | 4.32624 | 0.000049 |
| haPWV (m/s) | 0.8353 | 0.9627 | -4.96219 | 0.000005 |
| β-stiffness index from PWV | 2.4801 | 3.0110 | -2.92002 | 0.004688 |

H
| Variable | T-tests; Grouping: Carotid (Both in primary data.stw) |  |  |  |
| --- | --- | --- | --- | --- |
|  | Group 1: No<br>Group 2: Yes |  |  |  |
|  | Mean No | Mean Yes | t-value | p |
| Age | 49.3632 | 62.3779 | -6.47581 | 0.000000 |
| SBP rest | 117.9412 | 127.0000 | -2.50663 | 0.014480 |
| DBP rest | 76.9118 | 83.0256 | -2.31854 | 0.023304 |
| Body weight | 72.7735 | 81.1103 | -2.26866 | 0.026331 |
| BMI | 24.7749 | 28.6399 | -3.55828 | 0.000670 |
| Goal heart rate | 170.6368 | 157.6221 | 6.47581 | 0.000000 |
| Max HR | 152.2353 | 141.2308 | 3.42535 | 0.001024 |
| WT | 137.5000 | 114.1026 | 2.24521 | 0.027870 |
| METs | 7.7676 | 5.7692 | 4.71014 | 0.000012 |
| eVessel age | 47.5294 | 64.6154 | -6.85002 | 0.000000 |
| R-CAVI | 7.6265 | 8.7000 | -3.49087 | 0.000832 |
| L-CAVI | 7.6912 | 8.5872 | -3.06143 | 0.003109 |
| Mean SBP B | 127.3676 | 140.3974 | -4.60363 | 0.000018 |
| Mean DBP B | 80.7059 | 88.2308 | -4.22680 | 0.000069 |
| BP RB $(=(SBP+DBP)/2)$ | 97.9118 | 108.8718 | -4.34732 | 0.000045 |
| BP LB $(=(SBP+DBP)/2)$ | 99.8529 | 108.2564 | -3.51634 | 0.000767 |
| Mean BP B | 98.8824 | 108.5641 | -4.05994 | 0.000125 |
| BP RA $(=(SBP+DBP)/2)$ | 101.9412 | 112.8462 | -4.04464 | 0.000132 |
| BP LA $(=(SBP+DBP)/2)$ | 103.0000 | 113.2051 | -4.02214 | 0.000142 |
| Mean BP A | 102.4706 | 113.0256 | -4.27458 | 0.000059 |
| RTb | 86.6176 | 76.0256 | 3.55900 | 0.000669 |
| LTb | 81.5588 | 73.6154 | 2.57072 | 0.012246 |
| Mean Tb | 84.0882 | 74.8205 | 3.35620 | 0.001272 |
| Right Tba | 94.2059 | 78.6667 | 4.43953 | 0.000032 |
| Left Tba | 93.4118 | 79.8718 | 4.22990 | 0.000069 |
| Mean Tba | 93.8088 | 79.2692 | 4.37414 | 0.000041 |
| haPWV (m/s) | 0.8461 | 0.9674 | -4.68414 | 0.000013 |
| β-stiffness index from PWV | 2.5549 | 3.0133 | -2.48557 | 0.015291 |

I
| Variable | T-tests; Grouping: Brachiocephalic (Both in primary data.stw) |  |  |  |
| --- | --- | --- | --- | --- |
|  | Group 1: No<br>Group 2: Yes |  |  |  |
|  | Mean No | Mean Yes | t-value | p |
| Age | 50.1003 | 62.3642 | -5.92974 | 0.000000 |
| SBP rest | 118.3333 | 127.1081 | -2.42735 | 0.017748 |
| DBP rest | 76.6667 | 83.5946 | -2.66257 | 0.009586 |
| Body weight | 73.1472 | 81.1973 | -2.19058 | 0.031767 |
| BMI | 24.9034 | 28.7238 | -3.51942 | 0.000759 |
| Pulse rest | 66.5556 | 71.4865 | -2.38704 | 0.019651 |
| Goal heart rate | 169.8997 | 157.6358 | 5.92974 | 0.000000 |
| Max HR | 151.4722 | 141.3784 | 3.10945 | 0.002697 |
| WT | 136.1111 | 114.1892 | 2.09958 | 0.039321 |
| METs | 7.6444 | 5.7811 | 4.31716 | 0.000050 |
| Mean SBP B | 128.6111 | 139.8919 | -3.85507 | 0.000252 |
| Mean DBP B | 81.3889 | 87.9730 | -3.60347 | 0.000579 |
| BP RB $(=(SBP+DBP)/2)$ | 98.9444 | 108.4595 | -3.66602 | 0.000472 |
| BP LB $(=(SBP+DBP)/2)$ | 100.5000 | 108.0811 | -3.13005 | 0.002537 |
| Mean BP B | 99.7222 | 108.2703 | -3.50544 | 0.000794 |
| BP RA $(=(SBP+DBP)/2)$ | 102.9167 | 112.4865 | -3.46814 | 0.000894 |
| BP LA $(=(SBP+DBP)/2)$ | 103.5278 | 113.2432 | -3.79923 | 0.000303 |
| Mean BP A | 103.2222 | 112.8649 | -3.83495 | 0.000269 |
| RTb | 85.6111 | 76.4324 | 3.02549 | 0.003455 |
| LTb | 80.6111 | 74.1081 | 2.07795 | 0.041331 |
| Mean Tb | 83.1111 | 75.2703 | 2.78450 | 0.006867 |
| Right Tba | 92.5000 | 79.4865 | 3.58232 | 0.000620 |
| Left Tba | 92.3333 | 80.1892 | 3.71368 | 0.000403 |
| Mean Tba | 92.4167 | 79.8378 | 3.67203 | 0.000463 |
| haPWV (m/s) | 0.8603 | 0.9601 | -3.68383 | 0.000445 |
| β-stiffness index from PWV | 2.6092 | 2.9852 | -2.01532 | 0.047658 |

J
| Variable | T-tests; Grouping: Arterial Hypertension (Both in primary data.stw)<br>Group 1: No<br>Group 2: Yes |  |  |  |
| --- | --- | --- | --- | --- |
|  | Mean No | Mean Yes | t-value | p |
| Age | 51.4730 | 61.0834 | -4.52681 | 0.000021 |
| SBP rest | 117.8250 | 128.5000 | -3.27783 | 0.001564 |
| DBP rest | 77.7750 | 83.4500 | -2.32017 | 0.022947 |
| BMI | 25.6294 | 28.2240 | -2.44082 | 0.016921 |
| Goal heart rate | 168.5270 | 158.9166 | 4.52681 | 0.000021 |
| WT | 137.5000 | 113.7500 | 2.48550 | 0.015076 |
| METs | 7.4750 | 5.8675 | 3.95860 | 0.000165 |
| eVessel age | 49.3250 | 63.6750 | -5.57881 | 0.000000 |
| R-CAVI | 7.6525 | 8.7625 | -3.91296 | 0.000194 |
| L-CAVI | 7.7225 | 8.6400 | -3.35647 | 0.001222 |
| Mean SBP B | 128.4625 | 140.2875 | -4.50838 | 0.000023 |
| Mean DBP B | 82.8125 | 87.7375 | -2.74183 | 0.007574 |
| BP RB $(=(SBP+DBP)/2)$ | 99.0250 | 108.9250 | -4.22291 | 0.000065 |
| BP LB $(=(SBP+DBP)/2)$ | 101.3500 | 107.7250 | -2.82342 | 0.006028 |
| Mean BP B | 100.1875 | 108.3250 | -3.62953 | 0.000505 |
| BP RA $(=(SBP+DBP)/2)$ | 104.4750 | 112.3000 | -2.91720 | 0.004611 |
| BP LA $(=(SBP+DBP)/2)$ | 103.7500 | 113.5000 | -4.15705 | 0.000082 |
| Mean BP A | 104.1125 | 112.9000 | -3.69048 | 0.000413 |
| RTb | 84.8250 | 76.2250 | 3.00316 | 0.003590 |
| LTb | 80.7750 | 73.7500 | 2.30228 | 0.023987 |
| Mean Tb | 82.8000 | 74.9875 | 2.91367 | 0.004659 |
| Right Tba | 93.9250 | 78.6000 | 4.76792 | 0.000008 |
| Left Tba | 92.6750 | 80.1250 | 4.20660 | 0.000069 |
| Mean Tba | 93.3000 | 79.3625 | 4.53289 | 0.000021 |
| haPWV (m/s) | 0.8550 | 0.9692 | -4.63729 | 0.000014 |
| β-stiffness index from PWV | 2.6538 | 3.0123 | -2.00831 | 0.048072 |

K
| Variable | T-tests; Grouping: SCAD II-III (Both in primary data.stw)<br>Group 1: No<br>Group 2: Yes |  |  |  |
| --- | --- | --- | --- | --- |
|  | Mean No | Mean Yes | t-value | P |
| Age | 51.3686 | 74.1726 | -4.50496 | 0.000094 |
| Height | 172.5517 | 161.6667 | 2.14298 | 0.040345 |
| Pulse after stress | 86.3448 | 70.0000 | 3.15103 | 0.003673 |
| Goal heart rate | 168.6314 | 145.8274 | 4.50496 | 0.000094 |
| Max HR | 150.9655 | 170.3333 | -2.18173 | 0.037097 |
| Reached % | 89.6496 | 116.6267 | -4.95212 | 0.000027 |
| eVessel age | 50.1034 | 74.6667 | -3.35831 | 0.002146 |
| R-CAVI | 7.8897 | 11.2667 | -4.10892 | 0.000283 |
| L-CAVI | 7.9207 | 11.1333 | -4.10755 | 0.000284 |
| Right Tba | 94.0345 | 49.3333 | 5.57398 | 0.000005 |
| Left Tba | 93.3103 | 51.6667 | 5.70435 | 0.000003 |
| Mean Tba | 93.6724 | 50.5000 | 5.66552 | 0.000004 |
| Lha (cm) | 150.2887 | 141.4403 | 2.14298 | 0.040345 |
| haPWV (m/s) | 0.8723 | 1.1609 | -4.12256 | 0.000272 |
| β-stiffness index from PWV | 2.7431 | 4.4903 | -3.29422 | 0.002537 |
| eGFR (2021 CKD-EPI Creatinine) | 88.0345 | 67.4000 | 2.32528 | 0.027008 |

L
| Variable | T-tests; Grouping: Reason of discontinuation (Both in primary data.stw)<br>Group 1: Reach goal HR<br>Group 2: Horizontal ST depression >1mm |  |  |  |
| --- | --- | --- | --- | --- |
|  | Mean<br>Reach goal<br>HR | Mean<br>Horizontal ST depression<br>>1mm | t-value | P |
| Pulse after stress | 87.9014 | 77.4286 | 2.55692 | 0.012554 |
| R-CAVI | 8.0873 | 9.8286 | -3.40533 | 0.001058 |
| L-CAVI | 8.0873 | 9.5571 | -3.01437 | 0.003499 |
| Right Tba | 86.9859 | 73.1429 | 2.23774 | 0.028167 |
| Mean Tba | 86.9155 | 74.4286 | 2.13132 | 0.036296 |
| haPWV (m/s) | 0.9027 | 1.0327 | -2.75026 | 0.007439 |
| β-stiffness index from PWV | 2.7566 | 3.7569 | -3.27037 | 0.001616 |

M
| Variable | T-tests; Grouping: vessel stiffness (Both in primary data.stw)<br>Group 1: Normal<br>Group 2: High |  |  |  |
| --- | --- | --- | --- | --- |
|  | Mean<br>Normal | Mean<br>High | t-value | p |
| Age | 54.0870 | 59.0955 | -2.14345 | 0.035193 |
| SBP rest | 119.4667 | 127.9143 | -2.50815 | 0.014212 |
| Goal heart rate | 165.9130 | 160.9045 | 2.14345 | 0.035193 |
| Max HR | 149.8667 | 141.6000 | 2.70090 | 0.008479 |
| WT | 135.0000 | 113.5714 | 2.20767 | 0.030206 |
| METs | 7.1889 | 6.0057 | 2.76413 | 0.007119 |
| eVessel age | 50.0667 | 64.7714 | -5.71036 | 0.000000 |
| R-CAVI | 7.6156 | 8.9686 | -4.96292 | 0.000004 |
| L-CAVI | 7.6311 | 8.8886 | -4.87258 | 0.000006 |
| BP RA $(=(SBP+DBP)/2)$ | 105.6000 | 111.9714 | -2.31324 | 0.023345 |
| BP LA $(=(SBP+DBP)/2)$ | 106.0667 | 111.9143 | -2.31365 | 0.023322 |
| Mean BP A | 105.8333 | 111.9429 | -2.43662 | 0.017104 |
| RTb | 85.0889 | 74.6571 | 3.71174 | 0.000384 |
| Mean Tb | 82.2222 | 74.6143 | 2.80499 | 0.006349 |
| Right Tba | 90.7556 | 80.4857 | 2.94006 | 0.004316 |
| Left Tba | 90.5333 | 81.0857 | 2.99525 | 0.003674 |
| Mean Tba | 90.6444 | 80.7857 | 2.98776 | 0.003756 |
| haPWV (m/s) | 0.8652 | 0.9723 | -4.23338 | 0.000062 |
| β-stiffness index from PWV | 2.5951 | 3.1390 | -3.12731 | 0.002480 |

